# Neurons of the human subthalamic nucleus engage with local delta frequency processes during action cancellation

**DOI:** 10.1101/2024.12.02.24318298

**Authors:** Johanna Petra Szabó, Panna Hegedüs, Tamás Laszlovszky, László Halász, Gabriella Miklós, Bálint Király, György Perczel, Virág Bokodi, Lászlo Entz, István Ulbert, Gertrúd Tamás, Dániel Fabó, Loránd Erőss, Balázs Hangya

**Author notes:** Correspondence: Balázs Hangya.

## Abstract

The subthalamic nucleus (STN) is a key regulator of inhibitory control, implicated in decision making under conflict and impulsivity. Delta frequency oscillations, both in the STN and in frontal cortices have been associated with such active decision processes. However, it is yet unclear how neurons of the human STN are linked to local delta frequencies during response inhibition. Here, we recorded STN neurons and local field potentials (LFP) in human patients with Parkinson’s disease (PD) while they performed a stop-signal reaction time task during deep brain stimulation implantation surgery. Delta band LFP activity increased during stimulus processing in the STN. We found that half of the STN neurons responded to a diverse set of behaviorally relevant events that included go and stop signals, with a subset of neurons showing differential responses in successful and unsuccessful attempts at response cancelling. Failure to stop was associated with stronger go signal-related firing increase of STN neurons and their stronger coupling to local delta band LFP activity. Furthermore, a specific population of bursting STN neurons showed increased delta coupling. These suggest that the STN integrates go and stop signal-related information. Increased engagement of STN neurons with local delta band activity during stimulus processing impaired the ability to cancel the ongoing response. This effect may be linked to the disease-related rise in STN neuronal bursting. These findings may shed light on a potential neuronal mechanism linking cortical delta band processes with STN activity, both of which are critical elements in inhibitory control.

## Introduction

The hyperdirect pathway, connecting frontal cortical areas including the pre-supplementary motor area and the inferior frontal gyrus with the subthalamic nucleus of the basal ganglia, has been associated with inhibition of motor responses and canceling of actions^1–3^. More generally, it is deeply involved in determining when to act, impacting reaction time, impulsivity and decision making under conflict^4,5^.

Frontal cortical slow oscillatory activities were shown to be implicated in decision processes that involve inhibitory control^6–9^. Conflict anticipation, inferred from successful stopping of ongoing actions, was associated with a fronto-central power increase in the 3-5 Hz frequency band^7^. Low-frequency activity related to decision conflicts^10–12^ and specifically to action inhibition^13^ were also demonstrated in the STN. Importantly, the low-frequency synchronization of frontal cortex and STN reflected action cancellation and correlated with successful stopping^13^. However, how local neurons of the STN mediate this process has not yet been revealed, hindering a mechanistic understanding of conflict processing in the STN.

To address this, we recorded neuronal spiking activity from the STN while patients with Parkinson’s disease performed a stop signal reaction time (SSRT) task. This confirmed the presence of STN spiking related to go and stop cues^2,14^, but also demonstrated STN activity predictive of stop performance, violating former assumptions relegating such functions to more downstream circuits^2^, while more consistent with others assigning complex functions to STN including action planning^15,16^. Following up on this, we found that increased STN activity after go signals and stronger phase coupling to STN delta band activity hindered patients’ ability to stop.

PD is associated with an increase in bursting activity in the STN^17,18^. In accordance, we also observed that a large fraction of the recorded units exhibited bursting. A specific subset of strongly bursting units that showed suppressed activity after go signal presentation were more phase coupled to delta activity than weakly bursting units with similar response properties, suggesting that bursting may impair response inhibition via strong engagement with local delta band activity.

Deep brain stimulation (DBS) has been associated with reduced decision thresholds, leading to fast but inconsistent decisions^19,20^. We found that while this was indeed the case in some patients, other patients benefited from DBS surgery not only in their motor functions^21^ but also in their cognitive domain in the form of increased rather than decreased reaction times. These patients showed stronger STN bursting and signs of stronger engagement of STN units with local delta band processing, raising the possibility that patients with more severe STN dysfunction may benefit from DBS implantation with respect to their decision making.

## Results

### Stop Signal Reaction Time Task performed by patients with Parkinson’s disease

Patients with PD scheduled for DBS implantation surgery were involved in the study (Extended Data Tables 1 and 2). Patients were introduced to the task and performed the first behavioral test during preoperative evaluation on the day before surgery, after 12-hours withdrawal of PD medication (n = 13; right hand, n = 13/13; left hand, n = 11/13; preoperative test). Next, patients were tested during DBS surgery, while single and multiunit activity was measured from the STN. Thirteen patients participated in the intraoperative tests during which both the left hemisphere (buttons pressed by right hand) and the right hemisphere were tested in 12-12 patients. Finally, patients with PD enrolled in the study were tested approximately one year (13.1 ± 11.2 months, mean ± SD) post-surgery, after their optimal stimulation parameters were determined by the attending neurologist. The tests were performed after 12-hours withdrawal of PD medication, first for the left side (right hand), then for the right side (left hand) in combination with 60-channel EEG recording. Patients were first tested with their DBS stimulator turned off, then the test was repeated with stimulation turned on (DBS-off left hemisphere, n = 10; right hemisphere, n = 12; DBS-on left hemisphere, n = 10; right hemisphere, n = 5; postoperative test; Fig.1a).

**Figure 1.**
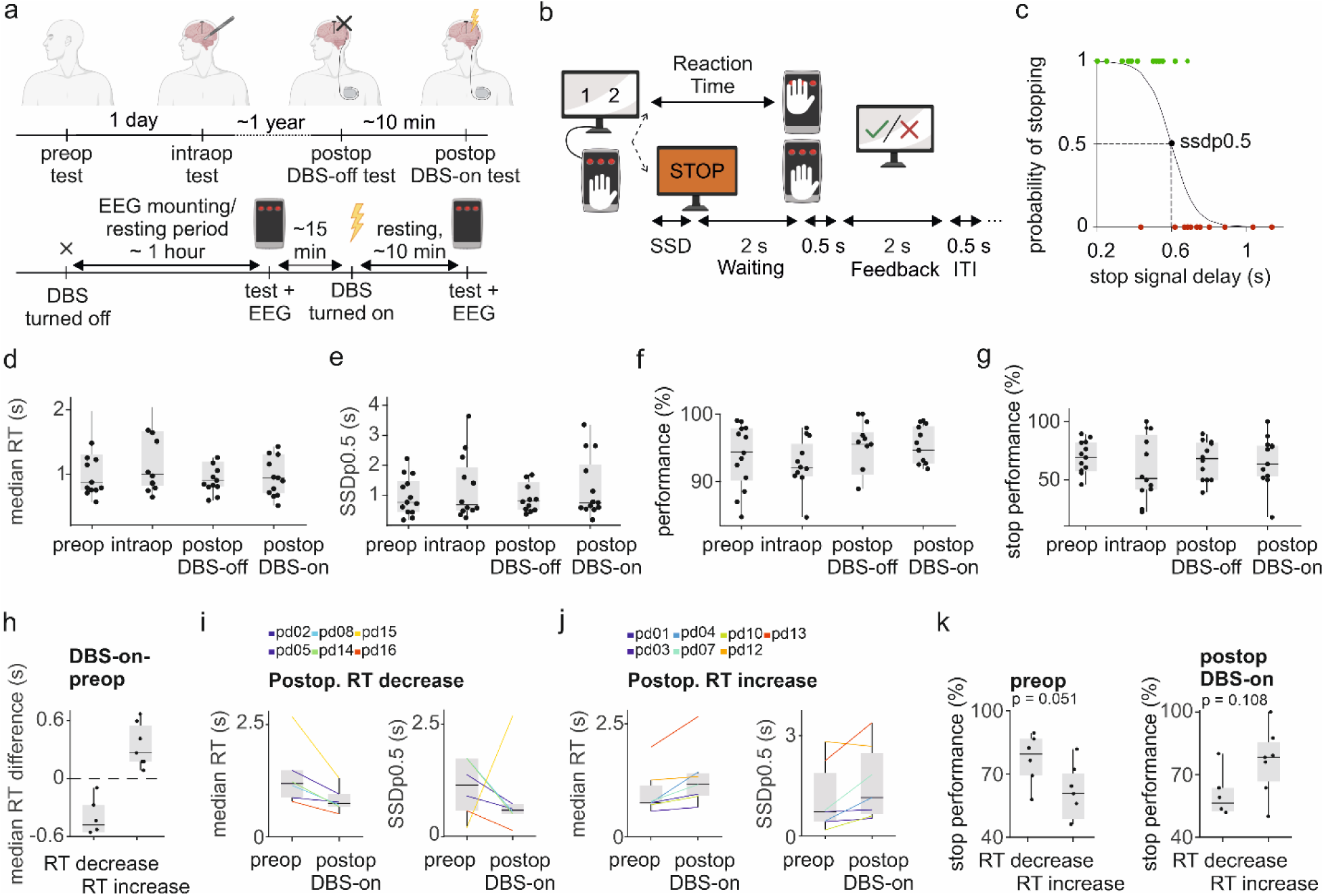
Stop Signal Reaction Time Task performed by patients with Parkinson’s disease. **a)** Timeline of the experiment**. b)** Schematic representation of the Stop Signal Reaction Time (SSRT) paradigm. Two numbers were shown on the screen simultaneously, next to each other (cue). Participants had to press the corresponding buttons (key press) in the correct order on a custom-designed button box. In a subset of trials, the cue was followed by a stop signal with variable delay (stop signal delay, SSD), when the key press had to be withheld. **c)** A generalized linear regression model was fitted to the SSDs corresponding to successful (green dots) and failed (red dots) stop trials to estimate the SSD with 50% probability of successful stopping (SSDp0.5). **d-g)** Median RTs (d), SSDp0.5 values (e), task performance (percentage of correct no-stop trials, f), stop performance (percentage of successful stop trials, g) in preoperative, intraoperative and postoperative experiments. Each dot represents the data of one patient (left side session). Box plots represent median, interquartile range, and non-outlier range. Outliers are not shown for better visualization; see Extended Data Fig. 1 for all data. **h-k)** Patients were grouped by the direction of RT change when comparing postoperative DBS-on to preoperative condition (left side sessions). **h)** Magnitude of RT change of patients with RT decrease and RT increase (left side). Outliers are not shown for better visualization; see Extended Data Fig. 2i-j for all data. **i)** RT (left) and SSDp0.5 (right) of patients with RT decrease. **j)** RT (left) and SSDp0.5 (right) of patients with RT increase. Each line corresponds to one patient. **k)** Stop performance in patients with RT decrease and RT increase, in preoperative (left) and postoperative, DBS-on measurements (right). Outliers are not shown for better visualization; see Extended Data Fig. 2g-h for all data.

To test inhibitory control in patients with PD, they performed a Stop Signal Reaction Time task. Patients viewed visual stimuli on a screen consisting of two integers from {1, 2, 3} presented next to each other, and they were instructed to press the corresponding buttons in the same order as quickly as they could on a custom-designed button box. After patient response, a feedback screen appeared stating whether the response was correct (over green background), or incorrect (over red background). In part of the trials, the screen featuring the stimuli was followed by a ‘STOP’ instruction presented at variable delays, prompting patients to withhold button pressing. If patients failed to withhold motor response, a feedback screen over red background informed them about the unsuccessful stopping. In case patients managed to refrain from pressing during a 2 second response window, a feedback screen over green background reinforced successful stop response (Fig.1b).

### Behavioral performance in the SSRT task

We measured patients’ reaction times (RT) and the stop signal delay (SSD): the estimated time delay between stimulus onset and stop instruction that resulted in 50% probability of successful response inhibition (SSDp0.5; Fig.1c). Both RT and SSDp0.5 showed considerable variation within and across patients and no systematic differences were seen in these parameters among pre-, intra- and postoperative tests including DBS-on and DBS-off measurements (Fig.1d-e), except for an intraoperative slowing in some of the patients that was significant when testing the right side (Extended Data Fig. 1e-f).

We also tested task performance (percent correct button presses) and stop performance (proportion of correct stops compared to all stop trials). While task performance was over 60% in 96 of 104 (92.3%) tests, it showed substantial heterogeneity without consistent changes from pre-to intra-to postoperative tests (Fig.1f-g), except for an intraoperative drop of performance that was significant for the right hemisphere tests (Extended Data Fig. 1g).

Although we did not find population level differences in SSRT performance related to DBS surgery or DBS stimulation, patients were categorized by either an increase or a decrease in RT following DBS surgery (DBS-on) compared to preoperative reaction times (Fig. 1h). As expected, SSDp0.5 estimations showed changes mostly concordant with median RT (Fig. 1i-j; Extended Data Fig. 2). Since average stop performance also changed in the direction of median RT, a better preoperative stop performance turned into worse performance postoperatively in the ‘RT decrease’ group (Fig. 1k), in agreement with a possible decrease in decision thresholds^19,22^.

Patients were categorized based on their clinical phenotype into tremor-dominated (TD; n = 5), akinetic-rigid (AR; n = 6) and mixed (MX; n = 2) groups. Tremor-dominated patients showed longer reaction times in preoperative tests and this difference appeared to be reduced after DBS surgery; however, these differences were not significant at this sample population size (Extended Data Fig. 3).

Unified Parkinson’s disease rating scale (UPDRS) scores, measuring PD severity, were determined both with (med-on) and without medication (med-off) preoperatively and postoperatively. These correlated positively with RT (significant for med-on in preoperative tests, both sides and for med-on and med-off postoperative DBS-on tests for the right side) and SSDp0.5 (significant for med-on and med-off postoperative DBS-on tests for both sides; Extended Data Fig. 4), suggesting that more advanced disease stages were associated with longer reaction times.

### Frontal and STN delta power increased during SSRT performance

Inhibitory control during response withholding involves top-down cortical processes^23, 24^. We sought for task-related changes in cortical processing during SSRT performance by aligning postoperative DBS-off EEG spectrograms to visual go signal and stop signal onsets (event-related spectrograms, ERS). We found an increase in delta band (1-4 Hz) power around go signals (Fig. 2a), which continued after stop signals in the stop trials (Fig. 2b), indicating increased frontal delta band activity during SSRT performance.

**Figure 2.**
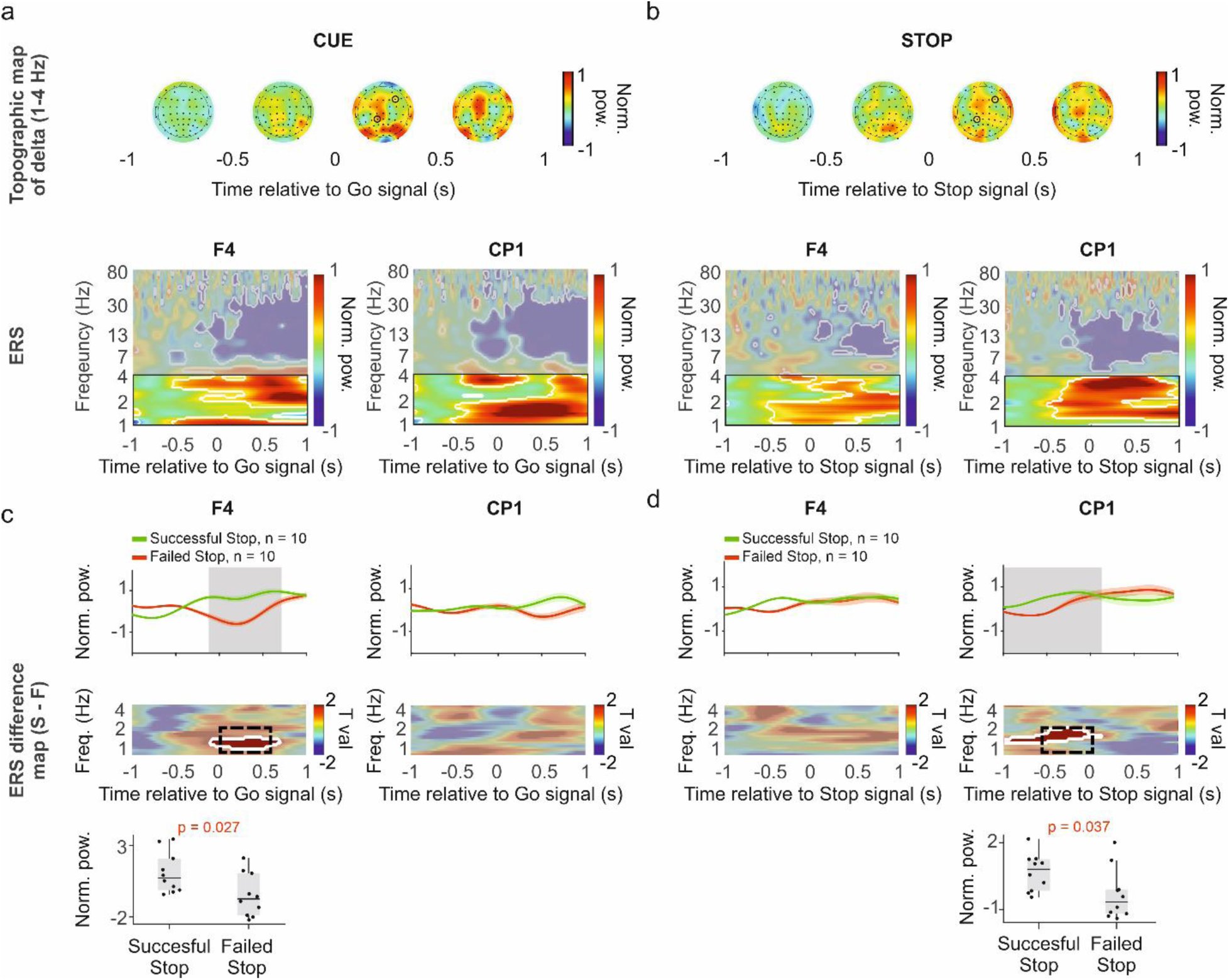
Frontal delta power increased during SSRT performance. **a**) Top, topographical maps (‘topo plots’) of 60 channel EEG recordings showing delta (1-4 Hz) power derived from scalp current densities, averaged across trials and patients in 0.5 s time windows aligned to go signals. Black dots correspond to the estimated position of EEG electrodes arranged according to standard 10-20 system. Data from left side postoperative DBS-off measurements are shown. Bottom, event-related spectrograms (ERS) of selected channels (marked by black circles on the topo plots) aligned to go signals (delta frequency range highlighted). Significant changes relative to baseline are indicated by white contours (permutation test with cluster-based correction, p < 0.05). **b)** The same as in panel a but aligned to stop signals. Frontal and centro-parietal delta power increase was detected both around go and stop signals. **c)** Top, delta band spectral power averages aligned to go signals for successful (green) and failed (red) stop trials, averaged across trials and patients from left side postoperative DBS-off measurements, shown for the same channels as in panel a (line and shading, mean ± SE). Significant differences are marked by grey shading (p < 0.05, permutation test with cluster-based correction). Middle, spectrograms displaying delta power differences between successful and failed stop trials, represented as paired T statistics (T val.) calculated for each time-frequency point. Significant changes relative to baseline are indicated by white contours (permutation test with cluster-based correction, p < 0.05). Bottom, delta power difference between successful and failed stop trials, calculated for the time-frequency windows marked in the middle panel (black dashed box). Box-whisker plots show median, interquartile range and non-outlier range. p = 0.027, Mann-Whitney U-test. **d)** The same as in panel c but aligned to stop signals. Successful stop trials were associated with larger delta power after the go cue and before the stop cue. p = 0.037, Mann-Whitney U-test.

When we compared stop trials in which the patient was successful in withholding response with unsuccessful stop attempts, we found that failed stops were associated with significantly lower right frontal (F4 channel) delta power after go signals (Fig. 2c) and lower left central (CP1 channel) delta power before stop signals (Fig. 2d). This may suggest that delta power increase in these regions contributes to better inhibitory motor control (Fig. 2).

Similar tendencies were observed with DBS stimulation turned on, although frontal-central delta increase was smaller in magnitude and shorter in duration and started earlier (Extended Data Fig. 5a-b), in line with a proposed reduction of inhibitory control during subthalamic stimulation^19^. Successful response inhibition was associated with higher delta band activity before the stop signal (C3 channel), consistent with DBS-off recordings (Extended Data Fig. 5c-d).

Having revealed delta band power changes in frontal EEG associated with the SSRT, we asked whether there were concomitant changes in delta band power in the STN local field potential (LFP). Therefore, we calculated event-triggered LFP averages (ETA, Fig. 3a, Extended Data Fig. 6a) and ERS of STN LFP power (Fig. 3b, Extended Data Fig. 6b) using the intraoperative microelectrode recordings performed during DBS surgery. We found that STN delta band power increased around go signal presentation (Fig. 3a,b), similar to what we found for frontal EEG channels in the postoperative DBS-off recordings. Moreover, this delta power increase was correlated with faster patient reactions, suggesting that go signal-related STN delta power increase facilitates cue detection (Fig. 3c). In contrast, stop signals were followed by a decrease in delta power (Extended Data Fig. 6b). We did not find significant differences in delta band power in the STN when we compared successful and unsuccessful stop attempts (Extended Data Fig. 6c).

**Figure 3.**
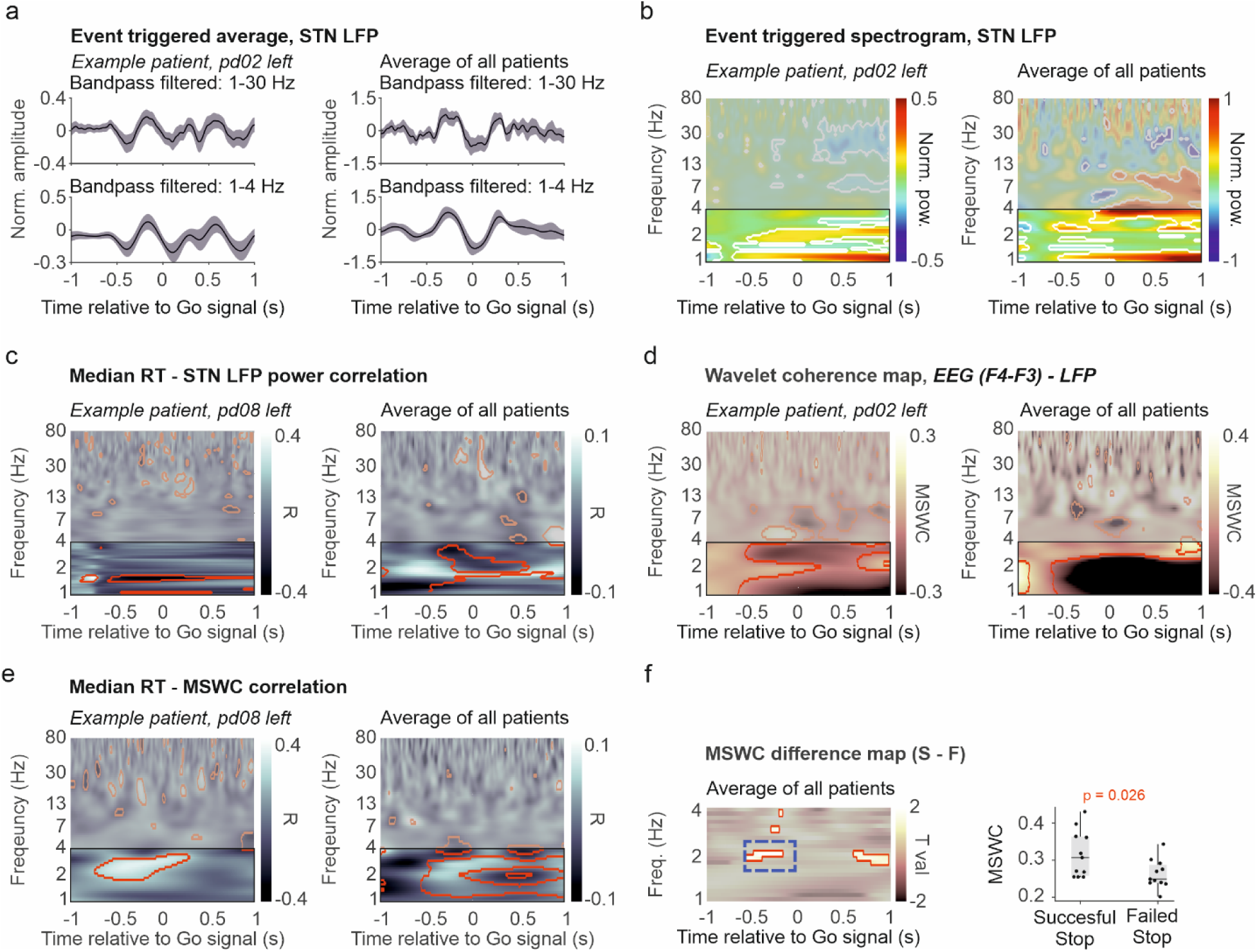
STN delta power increased during SSRT performance. **a)** Event-triggered averages (ETA) of STN local field potential (LFP) aligned to go signals, measured during DBS implantation. Broadband (top, 1-30 Hz) and delta band (bottom, 1-4 Hz) filtered signals are shown. Shaded lines mark standard error. Left, example patient; right, average across patients. **b)** Event-related wavelet power spectrograms (ERS) relative to baseline, aligned to go signals. Delta frequency band is highlighted; significant changes relative to baseline (−1 to −0.5 s) are marked by white contours. Left, example patient; right, average across patients. **c)** Time-frequency correlation of median RT and STN LFP power around the go signal (Pearson’s correlation coefficients). Left, example patient; red contours indicate regions with significant correlation calculated across trials (t-test, p < 0.05). Right, average across patients; red contours indicate significant changes in correlation relative to baseline (permutation test with false discovery rate correction, p < 0.05). **d)** Magnitude-squared wavelet coherence (MSWC) of STN LFP and intraoperative frontal bipolar EEG aligned to go signals. Color code indicates increase (white) and decrease (black) relative to baseline (−1 to −0.5 s). Delta frequency band is highlighted; significant changes relative to baseline are marked by red contours. Left, example patient; right, average across patients. **e)** Time-frequency correlation of median RT and MSWC around the go signal (Pearson’s correlation coefficients). Left, example patient; red contours indicate regions with significant correlation calculated across trials (t-test, p < 0.05). Right, average across patients; red contours indicate significant changes in correlation relative to baseline contoured (permutation test with false discovery rate correction, p < 0.05). **f)** Left, MSWC difference between successful and failed stop trials, aligned to go signals, represented as paired T statistics (T val.) calculated for each time-frequency point, averaged across patients. White color indicates greater coherence associated with successful stop trials (permutation test with cluster-based correction, p < 0.05). Significant differences are marked by red contours. Right, MSWC difference between successful and failed stop trials, calculated for the time-frequency windows marked in the left panel (blue dashed box). Box-whisker plots show median, interquartile range and non-outlier range. p = 0.026, Mann-Whitney U-test.

Next, we tested whether delta band activity changes were coordinated across frontal cortex and STN. For this, we used bipolar frontal EEG recordings obtained during DBS surgery. Interestingly, delta band coherence markedly decreased around go signal presentation (Fig. 3d) but increased right after stop signals (Extended Data Fig. 6d). Decreased delta coherence around go signals correlated with faster go responses (Fig. 3e). Nevertheless, successful response withholding was associated with higher frontal cortex-STN delta band coherence before (Fig. 3f), but lower after stop signals (Extended Data Fig. 6e). These results confirm that go signal detection and response do not rely on the hyperdirect pathway, in contrast with response inhibition^25^.

### STN neurons are responsive to behaviorally relevant events during SSRT

We recorded n = 193 units from the STN of 13 patients while they performed the SSRT task. Of these, n = 48 were considered single neuron activity based on standard cluster quality criteria (ID > 15 and L-ratio < 0.02), while the others were considered multiunits. Their localization within the STN was determined based on the postoperative reconstruction of the microelectrode tracks (Fig. 4a-e).

**Figure 4.**
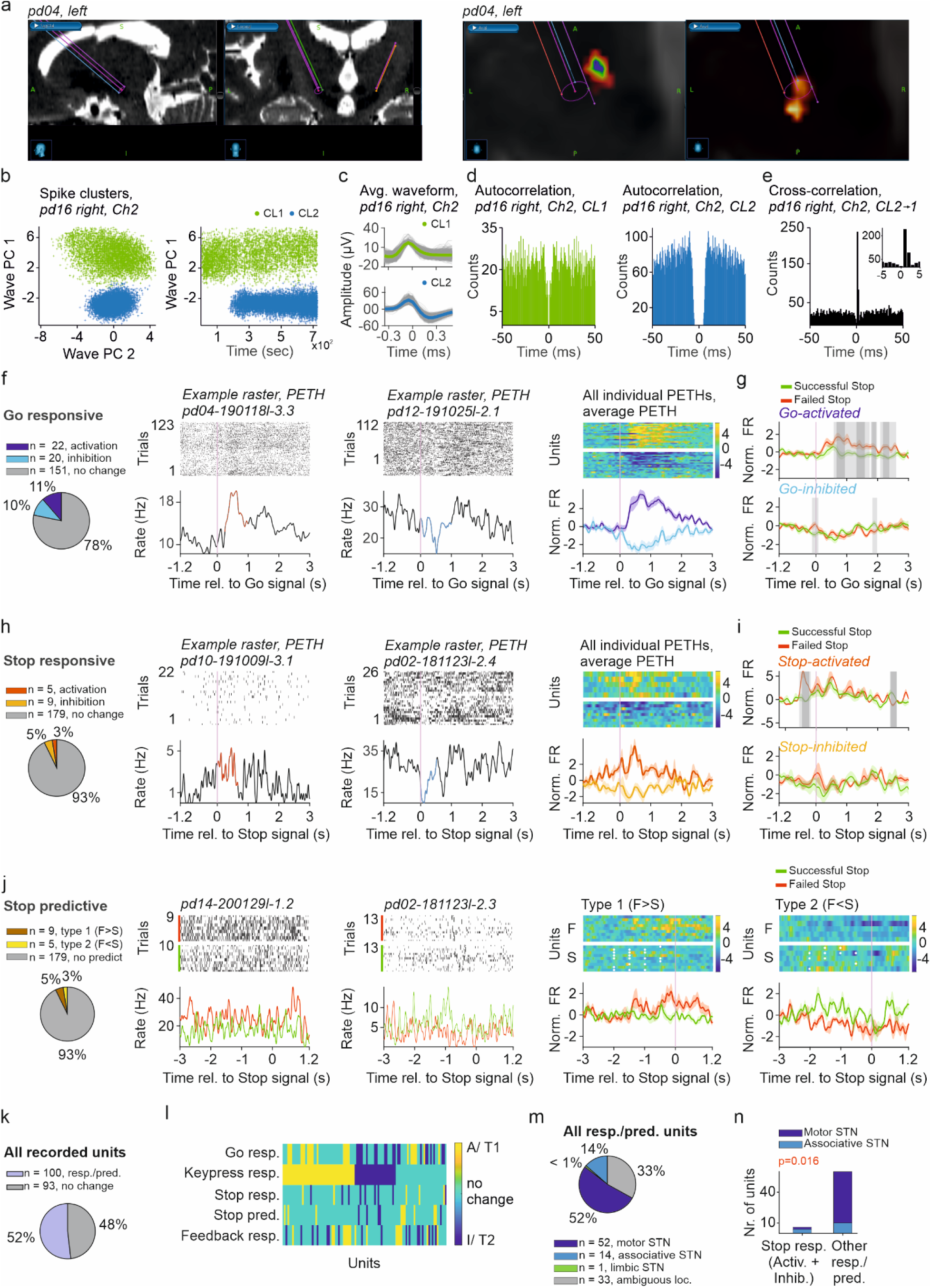
STN neurons are responsive to behaviorally relevant events during SSRT. **a)** Left, reconstruction of microelectrode leads of an example patient, based on preoperative MR and postoperative CT scans. Right, estimation of microelectrode position within STN subregions based on diffusion tensor imaging in the same example patient. Multicolor patches represent associative, red patches represent motor subregion. **b)** Clusters (CL) of spikes recorded by a microelectrode channel of an example patient. Left, spike clusters illustrated using first and second principal component of spike waveforms (Wave PC 1, 2). Right, Wave PC 1 as a function of time. Each color corresponds to a different cluster. **c)** Average waveform associated with clusters (CL1, top; CL2, bottom) shown in panel b. Individual spikes are shown in grey. **d)** Autocorrelograms corresponding to the example clusters (CL1, left; CL2, right) in panel b. **e)** Crosscorrelogram of the same clusters. Inset, middle values enlarged. **f)** STN units responsive to Go signals. Left, proportion of Go-responsive units. Middle, event-aligned raster plots and peri-event time histograms (PETH) for representative activated (significantly increased activity in red) and inhibited (significantly decreased activity in blue) units. Right, heatmaps of the Z-scored PETHs of all Go-responsive units (yellow, firing rate increase; blue, firing rate decrease) and average PETHs of activated and inhibited units (shading, standard error). **g)** PETHs of Go-activated and Go-inhibited units in successful (green) and failed (red) stop trials. Significant differences are marked by grey shading (light grey, p < 0.05; dark grey, p < 0.01, permutation test with cluster-based correction). **h)** STN units responsive to Stop signals. Same arrangement as in panel a. **i)** PETHs of Stop-activated and Stop-inhibited units. Same arrangement as in panel b. **j)** STN units predictive of stop trial outcome. Left, proportion of predictive units. Middle, event-aligned raster plots and PETHs for representative type 1 (lower activity preceding successful stop) and type 2 (higher activity preceding successful stop) units. Right, heatmaps of the Z-scored PETHs of all Stop predictive units (yellow, firing rate increase; blue, firing rate decrease) with average PETHs partitioned according to stop trial outcome (shading, standard error). **k)** Proportion of behaviorally responsive and/or predictive units. **l)** Activity change of responsive/predictive units to each behavioral event. Yellow, activated (A) and type 1 predictive (T1) units; blue, inhibited (I) and type 2 predictive (T2) units. **m)** Distribution of behaviorally responsive and/or predictive units across different STN subregions. **n)** Proportion of stop responsive (both activated and inhibited) and other types of responsive/predictive units in motor (dark blue) and associative (light blue) STN subregions. Stop responsive units were more frequently localized in the associative region (Fisher’s exact test, p = 0.016).

We found that many of these units responded to behaviorally relevant task events. First, we aligned spiking activity to go signals and calculated event-aligned raster plots and peri-event time histograms (PETH). These revealed that 22% of the recorded units changed their spike rate upon go signal presentation, with an equal portion of activated (n = 22, 11%) and inhibited (n = 20, 10%) units (Fig. 4f). When we separated the activity of go signal-responsive units in ‘stop’ trials based on whether the patient was successful in inhibiting the motor response, we found that failed attempts were preceded by a significantly larger go-elicited activation (Fig. 4g).

When we aligned unit activity to the stop signals, we found an 8% of units responding significantly to stop signal presentation: n = 5 (3%) of units were activated, while n = 9 (5%) showed inhibition (Fig. 4h). Although the percentage of significantly activated and inhibited neurons after stop signals was smaller than that after the go signals, this could also be due to the lower statistical power in the former analysis given that only a subset of trials were ‘stop trials’. At the same time, the average PETHs indicated firing rate changes already before the stop signals, suggesting that at least some of these units were affected by the go signals, with their firing rate changes continuing or accentuated after the stop signals. In line with this, ‘Stop-activated’ units showed higher firing rate for failed stop attempts before the stop signals (Fig. 4i), potentially corresponding to similar differences found for ‘Go-activated’ units (Fig. 4g).

Following up on this, we found units that showed differential activity before successful and unsuccessful motor inhibition, thus predicting the outcome of stopping (Fig. 4j). In n = 9 units (5%), activity was higher before failed stop attempts (type 1 predictive units), whereas in the case of n = 5 units (3%) we found the opposite, that is, faster spiking before successful motor inhibition (type 2 predictive units). Thus, type 1 predictive units were consistent with the differential responses to go signals in failed vs. successful stop trials shown above.

Next, we tested spike rate changes around key presses by aligning spike rasters and PETHs to the first button press of patients in each trial. We found that 36% of all units responded significantly to button press, with a dominance of activation (n = 44, 23%) over inhibition (n = 25, 13%; Extended Data Fig. 7a).

Finally, some of the recorded units (17%) responded to behavioral feedback and increased (n = 16, 8%) or decreased (n = 17, 9%) their firing upon presentation of the feedback screen (Extended Data Fig. 7b). However, upon closer inspection, the feedback-activated units seemed to show an activation that already started earlier and seamlessly continued after the feedback presentation, rather than a discrete increase at feedback onset.

Altogether, 52% (n = 100) of all units responded to at least one event (including predictive units), demonstrating an active engagement of the human STN during SSRT performance (Fig. 4k). The direction of go signal and key press responses were always concordant among units, whereas many STN units lacking significant go signal-elicited firing rate changes still responded to key presses, mostly by activation (Fig. 4l).

Most recorded units were localized to the motor part of STN (n = 85, 44%), with a smaller fraction in the associative part (n = 23, 12%) and only a few units were recorded in the limbic part (n = 3, 2%), whereas localization to identified parts of the STN nucleus could not be reliably carried out for n = 82 units (Extended Data Fig. 7c). Responsive or predictive units largely followed this distribution, with n = 52 units (52%) localized to the motor part of STN and n = 14 units (14%) localized to the associative part (Fig. 4m). Interestingly, units responding to the stop signals were more in the associative than in the motor STN, compared to other types of behaviorally relevant units (Fig. 4n; stop-responsive, 2/14 in motor, 4/14 in associative; other responsive/predictive, 50/86 in motor, 10/86 in associative; p = 0.0158, Fisher’s exact test). No other significant localization biases were detected (Extended Data Fig. 7d; Extended Data Table S3).

### STN neurons were phase coupled to local delta activity, especially before unsuccessful stop attempts

We demonstrated that when the patients were presented with the go signal, a significant fraction of STN units responded and, in parallel, delta power was increased in the STN LFP. We found that these two signatures of a change in activity were not independent, but a large fraction of units showed significant phase coupling (PC) to local delta band LFP activity both around the go (Fig. 5a-e) and the stop signals (Fig. 5f-j; Extended Data Fig. 8).

**Figure 5.**
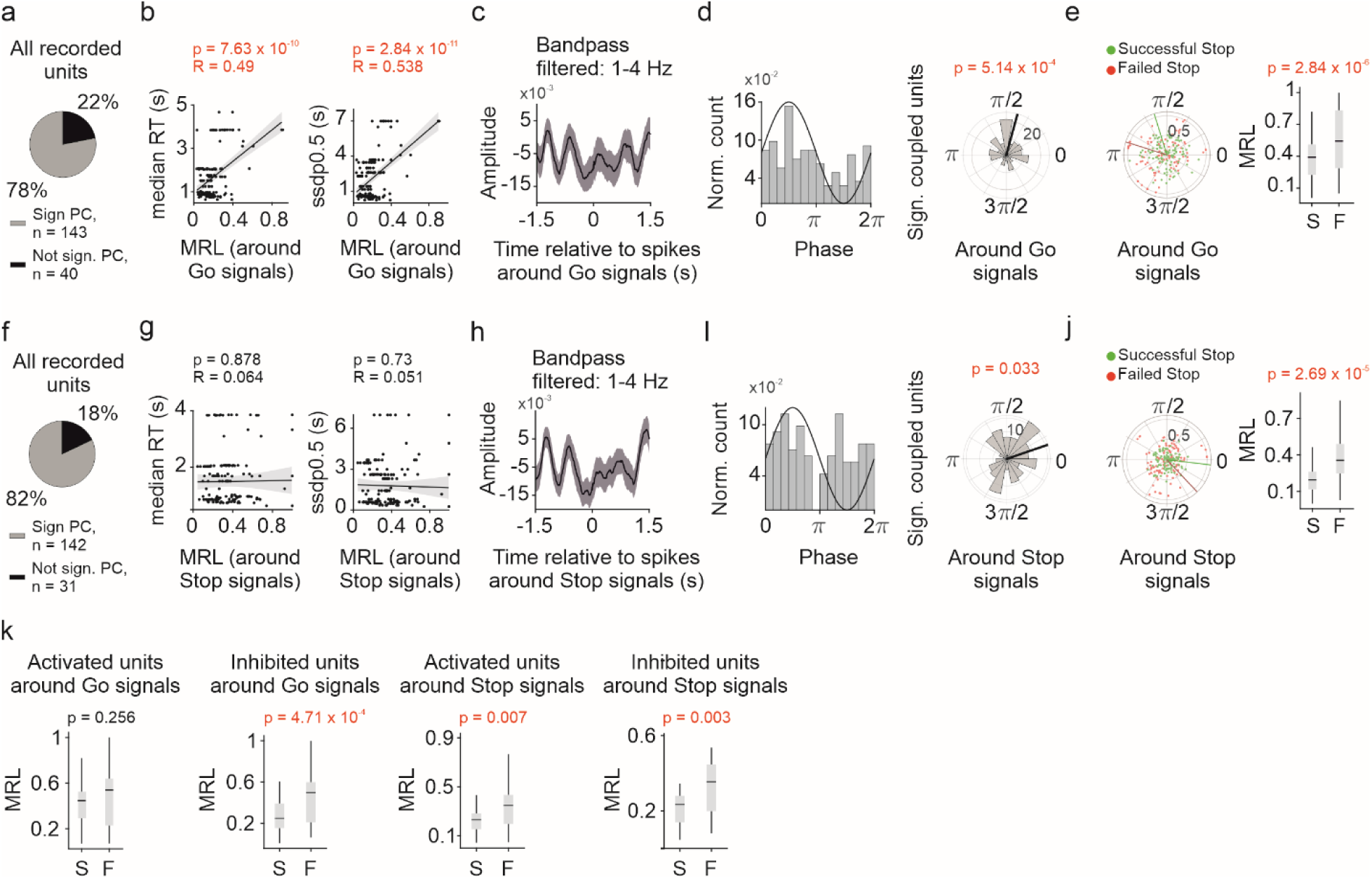
STN neurons were phase coupled to local delta activity, especially before unsuccessful stop attempts. **a)** Proportion of units significantly coupled to local delta oscillation from all recorded units. Phase coupling was calculated for a −1.5 1.5 s time-window around go signals. **b)** Correlation of phase-coupling strength (mean resultant vector length, MRL) around go signals and behavioral parameters: median RT (left) and SSDp0.5 (right). Each dot corresponds to one unit. Lines indicate robust regression with 95% confidence intervals (grey shading). Pearson’s correlation coefficients and corresponding p-values (t-test) are shown for each plot. **c)** Spike-triggered average of delta band (1-4 Hz) LFP, averaged across delta-coupled units for spikes −1.5 s to 1.5 s relative to go signals. Grey shading indicates standard error. **d)** Left, population phase histogram showing the distribution of preferred phases of delta-coupled units. Right, rose diagram of the same distribution. Circular uniformity was tested with Rayleigh’s test. **e)** Left, preferred phase and coupling strength (MRL) of delta-coupled units in stop trials, shown for successful (green) and failed (red) stop attempts. Right, comparison of coupling strength in successful (S) and failed (F) stop trials. Box-whisker plots show median, interquartile range and non-outlier range. p = 2.84 x 10^-6^, Mann-Whitney U-test. Outliers are not shown for better visualization; see Extended Data Fig. 8 for all data. **f-j)** Same as in panels a-f but calculated for spikes in a −1.5 s to 1.5 s time window around stop signals. **k)** Comparison of coupling strength between successful and failed stop trials in different behaviorally responsive populations (Mann-Whitney U-test).

We measured delta PC strength of STN units by calculating the mean resultant length (MRL^26^). The MRL around the go signal showed a strong positive correlation with both the median RT and the SSDp0.5 of the patients, showing that faster reactions were accompanied by weaker phase coupling (Fig. 5b). No such relationship was found for delta PC around the stop signals (Fig. 5g).

The recorded STN units showed delta phase preference also on the population level, most units being locked slightly before the peak of delta around the go signal (mean angle, 1.3 (74.5°); p = 0.0005, Rayleigh’s Z-test; Fig. 5c,d) and to the ascending slope around the stop signal (mean angle, 0.33 (18.9°); p = 0.0333, Rayleigh’s Z-test; Fig. 5h,i). The preferred phase was not significantly different in successful vs. unsuccessful stop trials, neither around cue (0.1 < p < 1, Watson’s two sample test of homogeneity; Fig. 5e), nor around stop signals (0.05 < p < 0.1, Watson’s two sample test of homogeneity; Fig. 5j). However, STN units showed stronger STN delta band LFP coupling associated with failed stop attempts, both measured around the go (p = 2.84 x 10^-6^, Mann-Whitney U-test; Fig. 5e) and the stop signal (p = 2.69 x 10^-5^, Mann-Whitney U-test; Fig. 5j). This difference was largest for units responding with inhibition to go signals (Fig. 5k).

Since we demonstrated frontal cortex – STN coherence in the delta band that was larger around the Go signals preceding successful stops (Fig.3d-f), we next tested if delta band phase locking could also be detected around the Go signals between STN units and frontal cortex EEG (Fig.6a).

**Figure 6.**
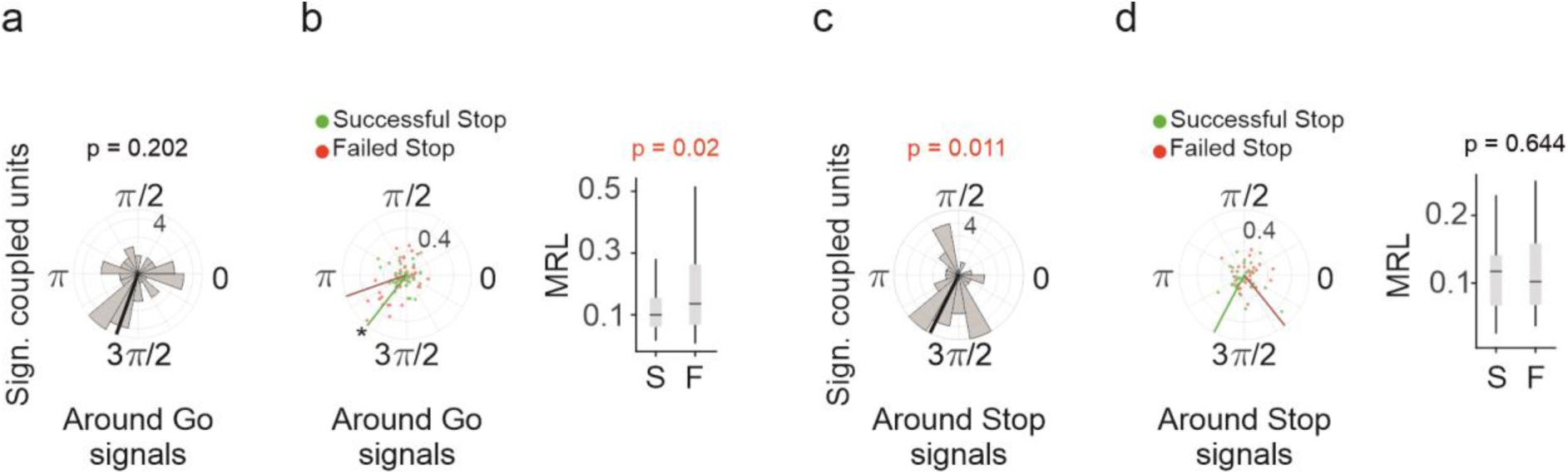
STN neurons showed significant population level phase locking to frontal delta before successful stops. **a)** Rose diagram of population phase histogram showing the distribution of preferred phases of units coupled to frontal delta, calculated for spikes in a −1.5 s to 1.5 s time window around go signals. **b)** Left, preferred phase and coupling strength (MRL) of frontal delta-coupled units in stop trials, shown for successful (S, green) and failed (F, red) stop attempts. Circular uniformity was tested with Rayleigh’s test (successful stops, p = 0.013; failed stops, p = 0.194). Right, comparison of coupling strength in successful (S) and failed (F) stop trials. Box-whisker plots show median, interquartile range and non-outlier range. p = 0.02, Mann-Whitney U-test. **c-d)** Same as in panels a-b but calculated for spikes in a −1.5 s to 1.5 s time window around stop signals. Circular uniformity was tested with Rayleigh’s test (successful stops, p = 0.277; failed stops, p = 0.57). Outliers are not shown for better visualization; see Extended Data Fig. 8 for all data.

We found a significant population level phase locking only before successful stops, suggesting a stronger frontal influence that could reflect hyperdirect pathway activity at the cellular level in the STN (Fig.6b). At the same time, individual STN neurons showed larger MRL and thus stronger phase locking before failed stops, similar to what was found locally in the STN. Neither of these differences were present around the stop signals (Fig.6c-d), again confirming the importance of the time window around the Go signal with respect to response inhibition.

### Bursting STN neurons are less responsive to cues, less predictive of inhibitory performance and a subset of them strongly lock to delta

Most STN units showed bursting activity (Fig. 7a), as expected based on previous literature^27,28^. We defined a burst index (BI) based on the presence of short-lag peaks (so called ‘burst shoulders’) in the spike autocorrelogram of units (see Methods^29^) and binarized units as bursting or non-bursting using an empirical BI threshold of 0.35 based on previous studies^30,31^. This resulted in 105 bursting (70%) and 45 non-bursting units (30%) out of 150 units with >1000 spikes. Average autocorrelograms confirmed the presence and absence of ‘burst shoulders’ in bursting and non-bursting units, respectively (Fig. 7b).

**Figure 7.**
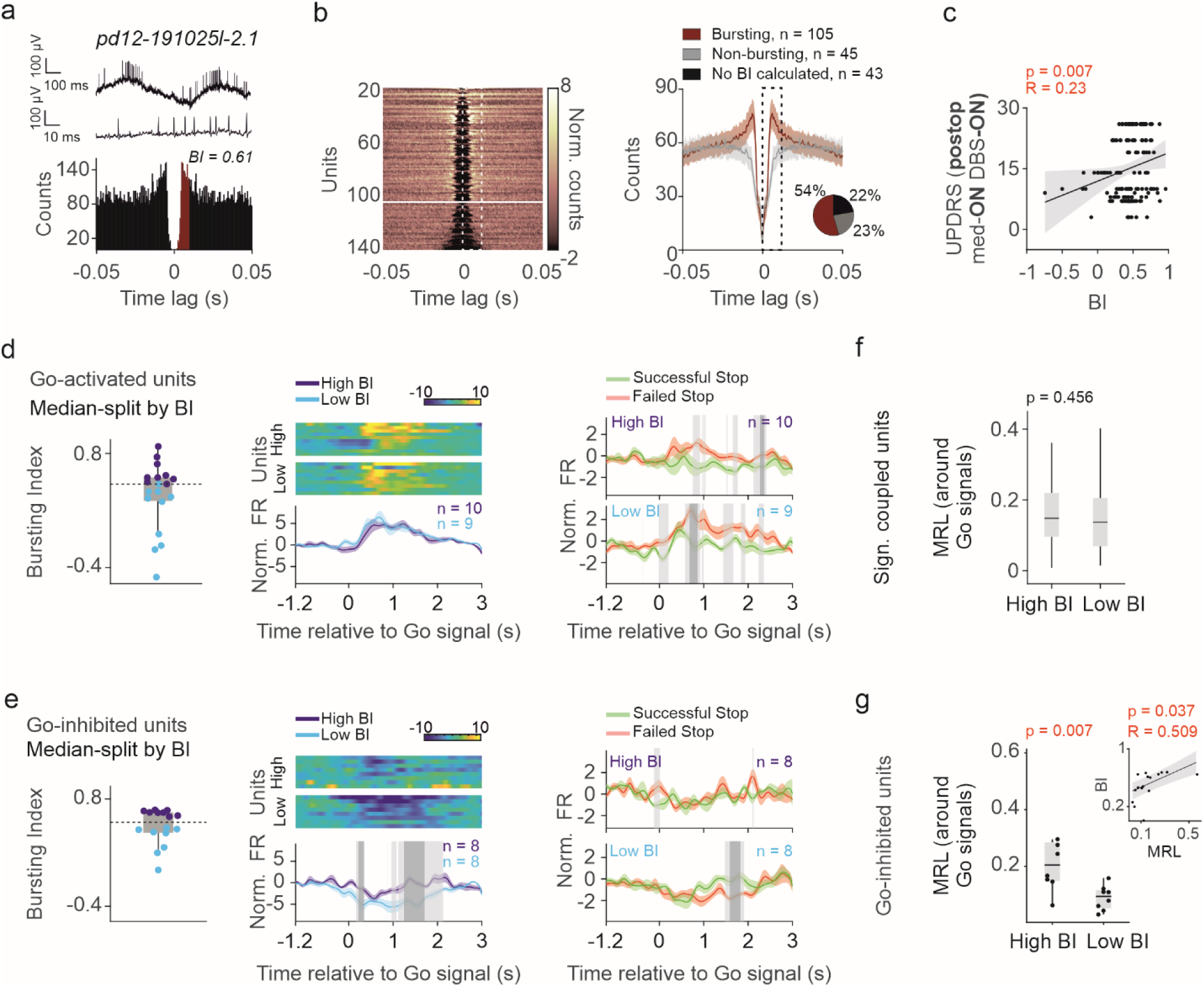
Bursting STN neurons are less responsive to cues, less predictive of inhibitory performance and a subset of them strongly lock to delta. **a)** Top, raw trace showing an example bursting unit. Bottom, spike autocorrelogram (AC) of the same unit with characteristic ‘burst shoulders’. The time lags from 0 to 10 ms used for calculating the bursting index (BI) are highlighted in brown. **b)** Left, ACs (color coded) of all recorded units with >1000 spikes. White dashed rectangle indicates the 0 to 10 ms time lags used for BI calculation. The white horizontal line marks the empirical BI threshold (0.35) separating bursting and non-bursting units. Right, average AC of bursting (brown) and non-bursting (grey) units; shading indicates standard error, black dashed rectangle indicates 0-10 ms time lags. The inset shows proportion of bursting and non-bursting units. **c)** Correlation of BI with postoperative UPDRS scores (medication on, DBS turned on). Each dot corresponds to a unit. Line indicates robust regression with 95% confidence intervals (grey shading). Pearson’s correlation coefficients and corresponding p-values (t-test) are shown. **d)** Go-activated units separated based on the median BI value. Left, box-whisker plot showing the BI value distribution. Dashed horizontal line indicates the median. Middle, peri-event time histogram (PETH) of each unit in the low BI and high BI groups (color coded; yellow, activation; blue, inhibition) and corresponding PETH averages (shading, standard error). Right, average PETH of successful (green) and failed (red) stop trials, for the low BI and high BI groups. Significant differences are marked by grey shading (light grey, p < 0.05; dark grey, p < 0.01, permutation test with cluster-based correction). **e)** The same as in panel d but for units inhibited in association with the go signals. **f)** Left, significantly delta-coupled units separated based on the median BI value. Delta coupling strength of low and high BI units calculated around go signals (−1.5 to 1.5 s time window) were not significantly different (p = 0.456, Mann-Whitney U-test). The box-whisker plot shows median, interquartile range and non-outlier range; see Extended Data Fig. 9f for all data. **g)** Go-inhibited units separated based on the median BI value. Delta coupling strength of low and high BI units were significantly different (p = 0.007, Mann-Whitney U-test). Each dot corresponds to a unit. The inset shows correlation between MRL around go signals and BI of go-inhibited units. Line indicates robust regression with 95% confidence intervals (grey shading). Pearson’s correlation coefficients and corresponding p-values (t-test) are shown.

In line with previous studies, bursting was stronger towards the dorsal part of STN and closer to the centroid of the motor subregion^32^ (Extended Data Fig. 9a-c). Postoperative UPDRS scores (DBS stimulator turned on) showed a significant positive correlation with the BI of the recorded units (Fig. 7c). Moreover, postoperative UPDRS scores correlated better with STN bursting than preoperative UPDRS scores (Extended Data Fig. 9d), which may indicate that motor symptoms in DBS-implanted patients have a strong connection with STN function. We did not find any significant correlations between median RT or SSDp05 and BI (Extended Data Fig. 9e).

Next, we asked whether STN units responsive to go signals differed in their activities depending on their burstiness. By performing a median split of STN units based on BI, we found that strongly and weakly bursting go-activated STN neurons showed comparable responses to the go cues. When we compared successful and failed stop trials, stronger go-elicited activation was associated with failed stops in both groups (Fig. 7d), similar to what we found when all units were considered (see Fig. 4b). Nevertheless, the difference was larger and appeared earlier for weakly bursting units.

We performed the same analysis for units that were inhibited following the go signals. Here, we found that weakly bursting neurons were characterized by significantly larger inhibitory responses than strongly bursting units. These weakly bursting units also showed larger go signal-associated inhibition in failed vs. successful trials (Fig. 7e). Thus, strongly bursting units were both less responsive and less predictive of inhibitory control performance, likely representing a Parkinson-related STN dysfunction.

Strongly bursting units coupled to a significantly later phase of STN LFP delta activity (mean angle, 2.44 (139.8°) for high BI and 0.77 (44.1°) for low BI; p < 0.05 Watson’s two sample test of homogeneity, Extended Data Fig. 9f), with similar phase coupling strength as weakly bursting units (Fig. 7f, Extended Data Fig. 9f), when examining phase-coupling around go signal presentation (but not around stop signals, Extended Data Fig. 9g). When we focused on go-inhibited neurons where the largest difference between weakly and strongly bursting neurons was seen (Fig. 7e), we found stronger delta PC in strongly bursting neurons as well as a positive correlation between BI and PC strength measured by MRL (Fig. 7g, Extended Data Fig. 9i). Since stronger delta phase locking was associated with unsuccessful response inhibition (Fig. 5e and j), it is conceivable that the disease-related dominance of bursting STN units in patients with PD impairs inhibitory control.

### More severe STN dysfunction at the recording site predicts RT increase after DBS surgery

About half of the patients showed a postoperative decrease of RTs, which could be a sign of decreased decision thresholds according to previous reports^33^, whereas other patients showed a postoperative RT increase (Fig. 1h-k). UPDRS scores, year of disease onset, disease duration, indices of postoperative motor improvement and levodopa equivalent daily dose (LEDD) reduction were assessed, but neither of these explained this difference (Extended Data Fig. 10a-c).

We tested whether STN activity was predictive of this outcome of the interventions. Interestingly, we found that STN units recorded from patients with RT increase showed higher burst index (Fig. 8a). It is conceivable that small differences in electrode positioning could underlie this effect, that is, targeting a region with more bursting neurons more likely led to an increase of RT. However, when we carried out a systematic analysis of potential correlations of stimulating electrode positioning and postoperative reaction times, we only found relatively weak evidence for this (Extended Data Fig. 10g-j). Alternatively, the STN of patients with postoperative RT increase could be more severely affected by the disease, reflected in more pathological bursting. Since an RT increase could indicate increased decision thresholds and reduced impulsivity after DBS implantation, more pathological bursting in the recording area could foreshadow a better DBS intervention outcome in the decision-making domain.

**Figure 8.**
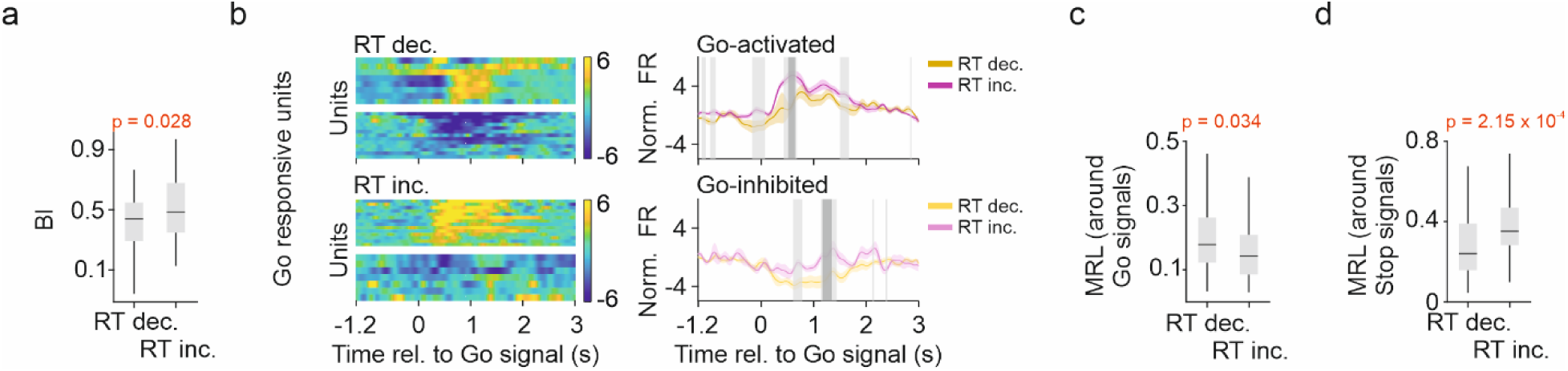
More severe STN dysfunction at the implantation site predicts RT increase after DBS surgery. **a)** Comparison of BI of STN units recorded from patients with RT decrease (RT dec.) and RT increase (RT inc.; p = 0.028, Mann-Whitney U-test). Box plots represent median, interquartile range, and non-outlier range. Outliers are not shown for better visualization; see Extended Data Fig. 10d for all data. **b)** Left, peri-event time-histogram (PETH) of go-responsive units (color coded; yellow, activation; blue, inhibition). Right, corresponding PETH averages. Significant differences between PETHs of patients with RT decrease and RT increase are marked by grey shading (light grey, p < 0.05; dark grey, p < 0.01, permutation test with cluster-based correction). **c)** Comparison of delta coupling strength of STN units recorded from patients with RT decrease vs. RT increase (spikes in −1.5 s to 1.5 s time window around go signals were included; Mann-Whitney U-test). Box plots represent median, interquartile range, and non-outlier range. Outliers are not shown for better visualization; see Extended Data Fig. 10n-o for all data. **d)** Same as in panel c but calculated for spikes in a −1.5 s to 1.5 s time window around stop signals.

We also found that STN units recorded from patients with RT increase showed larger activation after go signals (Fig. 8b), which was associated with failed stop attempts when all responsive units were considered (see Fig. 4b), suggestive of a more severe STN dysfunction at the recording site. STN neurons inhibited around go signals showed weaker inhibition in patients with postoperative RT increase (Fig. 8b), a possible consequence of stronger bursting activity that was associated with smaller responses in this group of units (see Fig. 7f; no difference in the response magnitude of stop-responsive units were found, Extended Data Fig. 10l).

The number of delta-coupled units (Extended Data Fig. 10m) and the mean phase of coupling around go and stop signals were similar in the two patient groups (Watson’s two sample test of homogeneity, p > 0.05; Extended Data Fig. 10n-o). Interestingly, STN units in patients with RT increase were less coupled to delta activity around go signals (Fig. 8c), but more strongly coupled around stop signals (Fig. 8d). The latter difference was larger, again suggesting a stronger local STN impairment in patients with postoperative RT increase, based on the association of stronger delta coupling with failed stop attempts (see Fig. 5e,j).

## Discussion

### Stop Signal Reaction Time Task before, during and after DBS implantation surgery

We have examined the SSRT performance of 13 patients with PD prior to, during and following DBS implantation surgery. We have not observed consistent changes in behavioral measures on a population level; however, there were significant changes on the level of individual patients: RT either increased or decreased following surgery compared to preoperative tests.

Numerous conflicting findings were reported on the effect of STN DBS on response control. While some researchers found unchanged RT with improved SSRT in response to stimulation^34,35^, others described faster RTs along with unaffected^36^, prolonged^37,38^ or even improved SSRT^39^. A possible reason underlying these inconsistencies is that the STN is involved in distinct processes of inhibitory control. On one hand, it provokes an abrupt movement suppression (reactive stopping)^40^, triggered either by a stop signal, canceling the already initiated response or by a conflict-inducing stimulus, calling for prolonged evidence accumulation^40^. In accordance, lesion studies of the STN showed impaired ability to inhibit initiated responses^41^. On the other hand, STN was suggested to control the tonic restraint (proactive inhibition) maintained to prevent premature responses^42^. Similarly to reactive stopping, proactive inhibition presumes increased decision bound in face of conflicting situations, however, in a strategic manner, preceding stimulus presentation. Greater RT difference between low- and high-conflict trials observed during stimulation^38^ suggested a beneficial effect of STN DBS treatment on the control of proactive inhibition. Similar conclusions were drawn by manipulating bilateral STN DBS in a countermanding task^42^. Additionally, despite the increase in impulsive errors produced by STN stimulation^19,33,40^, it simultaneously enhanced top-down suppression of conflict-induced interference effects^43^. Therefore, while decision thresholds are thought to decrease during STN DBS therapy, the better top-down control of proactive inhibition might partially counteract this effect resulting in reduced, but more dynamic threshold adjustments. Unilateral DBS applications in some studies may show limited effectiveness compared to bilateral implants, representing an added layer of variance across studies^44^.

Reaction times might also be influenced by impaired motor execution, which may be reflected by the generally positive correlations between UPDRS scores and reaction times. In this regard, shorter RT’s might not only reflect impulsive decisions but also improved motor execution^45^. An analysis of performance did not resolve this question due to low error rates and variability in behavior. Additionally, STN may also modulate non-motor functions, such as discriminative attention^46^ or regulation of emotional relevance^47,48^, which further complicates the interpretation of stimulation effects. However, altogether, the group of patients demonstrating slower RT following DBS implantation might have improved in proactively inhibiting their movements^40^. Importantly, the changes in RTs were concordant between DBS-off and DBS-on conditions, indicating a long-term stimulation effect. In contrast with the prompt diminution of motor symptoms in response to stimulation, cognitive effects may emerge more gradually over the long-term treatment.

### Frontal and STN delta power increased during SSRT performance

We found increased delta band activity around go signals both in the frontal cortex and in the STN^49^, and stronger STN delta power around go signals was associated with faster patient reactions, as also shown before^50–52^. In the frontal cortex, delta activity continued to increase after stop cues, whereas the STN showed a concurrent decrease. These suggest that STN delta may be more related to go signal processing, while frontal delta rather correlates with conflict anticipation^50,53^ and inhibitory control^12^. Consistent with this, albeit frontal-STN delta band coherence decreased during the go signal, maintaining a relatively higher coherence was a prerequisite of successful stopping. Failed stop attempts were associated with lower fronto-central delta power that could be a sign of decreased top-down influence through the hyperdirect pathway^25^ and were followed by an increase of frontal-STN delta band coherence, possibly indicating top-down control signals arriving late^2^.

When comparing EEG recordings with DBS stimulation off and on, we found that the frontal delta band power increase related to go signal presentation was smaller in magnitude and shorter in duration with DBS turned on. This is consistent with a reduction in inhibitory control during STN stimulation, as earlier reports proposed^20,54^. Higher frontal delta band activity before stop signals were associated with successful response inhibition both DBS on and off, emphasizing the role of the hyperdirect pathway during a brief time window around the stop signal in inhibitory motor control^55^.

Cavanagh et al. proposed that slow frequency bands may reflect a neural substrate for cortico-basal ganglia communication regulating decision processes^12^, whereas in another study Tan and Brown found that force and movement vigor were rather represented by higher frequency bands in the STN^56^. Our results largely confirm this; however, we found that STN delta band activity around the go signals may partly represent go stimulus processing, also confirmed by the association of larger STN delta power increase with faster patient responses to go signals. Concomitantly, lower frontal-STN delta coherence was also associated with faster RTs, further supporting the inhibitory role of STN-frontal synchronization in the delta band. These observations are supported by previous reports linking faster RTs to larger STN power and lower frontocentral-STN phase-locking in the low frequency range^57^. This makes the STN an integration site of go and stop processing, resonating with the race model of action cancelation proposed by Schmidt and Berke^2^, but suggesting that integration not only happens in the substantia nigra pars reticulata, but already upstream in the STN, consistent with the STN being a key hub for decision making under conflict^58^.

### Increased go signal-related activity and local delta band synchrony in the STN impairs inhibitory control

Although a number of studies investigated STN LFP and also delta band activity in particular, as discussed above, few animal^2,59^ and even scarcer human studies^60^ investigated the neuronal activity in the STN with respect to inhibitory motor control^14,15,25,61,62^ or decision making under conflict^63–65^. Therefore, the relation of human STN spiking with local delta band activity remained unclear. We found that about half of the STN units exhibited diverse responses to behaviorally relevant events including go signals^59^, stop signals^2,14^ and key press movements^14^, demonstrating an active engagement of the STN during SSRT performance, in line with previous animal and human recordings.

Consistent with the finding that STN delta band activity might at least in part reflect go instruction processing, failed attempts to stop were associated with increased go-signal-evoked activation in the STN, potentially indicating more advanced cue processing hindering the ability to stop. This is supported by reports on the valence specificity of STN neurons responsive to both go and no-go stimuli^61^. In line with this, a subset of neurons differentiated successful from failed stop attempts by differential firing rates before stop signals, thereby predicting inhibition outcome. While some animal^2^ and human studies^14^ found fast stop-signal-elicited responses in STN neurons emphasizing its role in reactive stopping, the presence of outcome predictive STN units in comparable proportions supports the notion of STN being also involved in proactive inhibition^66^. Combining this with former reports^14,67^, it is conceivable that the ventromedial STN is more associated with reactive stopping, while proactive inhibition is localized more to the dorsolateral part. A larger sample of unequivocally localized STN units will be required to determine this in the future.

We found that local STN delta power and unit activity were not independent: a large fraction of STN units showed phase coupling in the delta frequency band with most units preferably locked to the rising phase of delta. Somewhat unexpectedly, stronger phase locking predicted slower patient responses to go signals, suggesting that a strong delta phase coupling was not beneficial to go instruction processing. Alternatively, slower RTs might have reflected, at least in part, proactively inhibited responses. In line with this, a subset of STN units were reported to couple more strongly to low frequency oscillations (2-8 Hz) during high conflict trials, with similar phase preference as shown by our data^64^. However, despite longer reaction times, stronger phase locking was also associated with failed stop attempts. This association was particularly apparent for STN units that were inhibited upon go signals, suggesting that this group might be of special relevance in response inhibition. Although the presence of such neurons was reported in monkeys (Fig. 3 in ref. ^59^), their roles in neuronal processing and control of behavior have not yet been investigated. The special role of go-inhibited units in cognition was also highlighted by a study on speech production, where STN units demonstrating decreased firing rate were linked to cognitive aspects of speech, while those producing increased firing rate were associated with motor aspects.^68^ Thus, increased local delta band synchrony, indexed by stronger phase coupling of STN units to local delta band activity, might impair reactive inhibitory motor control. This supports the hypothesis that the ability to release proactive inhibitory control is impaired in PD patients^66^, hampering both movement initiation and switching to reactive stopping.

Bursting activity of STN neurons was found to be increased in PD; therefore, it is considered part of the pathophysiology^17,18^. However, patients with essential tremor also showed STN bursting in a smaller fraction of cells, suggesting that PD only enhances bursting activity that may normally also be present^18^. Our estimates of the proportions of bursting STN units in PD patients matched former reports^18^. Moreover, STN bursting exhibited a significant positive correlation with the postoperative UPDRS score with DBS turned on, suggesting that the presence of many strongly bursting STN units predicted worse postoperative motor outcome. This association of bursting with postoperative UPDRS evaluation surpassed that of with preoperative UPDRS scores, suggesting that clinical symptoms in DBS-implanted patients have a strong connection with STN function.

STN units inhibited upon go signals with less bursting showed larger inhibition, especially in failed stop trials. This mirrors the larger activation of go-signal-activated units in failed stop attempts, raising the possibility that local inhibitory connections shape the activity of these neuronal groups^69^. In the same group of go-inhibited units, more bursting was associated with stronger delta phase locking, which we found detrimental to SSRT behavior (see above). This was further substantiated by a significant positive correlation between bursting strength and delta band phase locking. Altogether, stronger bursting was linked to weakened responsiveness, stronger delta phase locking and less predictive power with respect to movement cancellation performance, suggesting that the dominance of the bursting STN phenotype in patients with PD impairs inhibitory motor control.

### Stronger burst firing activity at the implantation site predicted a postoperative increase of reaction times

The sampled STN region of patients showing increased RT following STN implantation proved to be more affected by the disease, indicated by higher bursting index of STN units. Bursting especially affected the activity of go-inhibited units in this patient group that showed diminished responsiveness. At the same time, greater responsiveness of go-activated units, associated with unsuccessful stops might represent stronger impairment of inhibitory control. Larger go-activation due to better movement initiation was not likely, since the preoperative RT of patients with postoperative RT increase and RT decrease was comparable.

In line with this, there is evidence that patients with STN neurons demonstrating higher intra-burst frequency, i.e. more pathologic STN functioning at baseline were less prone to develop impulsive-compulsive behaviors following STN DBS implantation, regardless of motor symptoms^70^. Another study found that stopping functions might be altered in opposite directions depending on baseline performance: stimulation improved response inhibition in patients with slower baseline SSRT compared to control subjects, while impaired stopping in those with baseline scores matching the control group^36^. This supports the idea that increased RT observed in our data is associated with better control of proactive inhibition.

The detrimental effect of burst firing on the responsiveness of STN units was more accentuated in go-inhibited units. This raises the possibility that the direction of change exerted by stimulation on inhibitory control depends on the integrity of go-inhibited units.

Since bursting index correlated positively with delta phase locking strength, greater coupling would be expected in patients with RT increase, which was found around stop signals but not around go signals. Nevertheless, the delta phase locking difference was larger in association with the stop cues, suggesting stronger local delta synchrony of STN units in patients with RT increase. Provided that stronger STN delta phase locking correlated with failed stop attempts, this may also indicate the STN’s involvement in impaired decision making in the patient group where the implantation surgery led to slowed behavioral responses.

It remains open whether the differential STN activity patterns we observed that predicted the effects of DBS on RTs reflected a general difference across patients with respect to their STN dysfunction, differential STN firing patterns at the recorded area influenced by differential electrode positioning, or a combination of both. In support for the role of the position of the stimulated contact, we found that patients with faster RTs were stimulated closer to the limbic centroid of the right STN. This would imply that with carefully chosen locations for stimulation, a better outcome on impulsive decisions could be achieved; however, further specific tests of this hypothesis will be required. Nonetheless, this notion is supported by former studies, which found that stimulation of ventral STN caused inhibitory deficit^71^, while stimulation of dorsal STN improved inhibitory functions^72^. While early implantation was shown to result in motor improvements comparable to later stage surgeries^73,74^, local STN pathology has not been assessed in these studies.

## Materials and Methods

### Patient selection

Thirteen patients with Parkinson-disease (6 males; age, 60.92 ± 12.6 years; Extended Data Table 1) undergoing bilateral STN DBS implantation (Extended Data Table 2) were enrolled in this study, recruited from the Clinic for Neurosurgery and Neurointervention (formerly National Institute of Clinical Neuroscience) of Semmelweis University (Budapest, Hungary). Informed consent was obtained from all subjects. Informed consent forms and all research protocols were approved by the Institutional Research Ethics Committee of NIMNN or its predecessors (OKITI IKEB1/2018; OMIII IKEB 21/2023) and the National Scientific and Ethical Committee of the Medical Research Council, Budapest, Hungary (OGYÉI/64814-6/2023). All research methods were performed in accordance with the relevant guidelines and regulations (Clinical Trials Directive (EC) No. 2001/20/EC, Note for Guidance on Good Clinical Practice (CPMP/ICH/135/95), the Declaration of Helsinki, Codex of Bioethics (2nd edition, Medical Research Council, Hungary), and Hungarian regulations Act CLIV of 1997 on Health; Act C of 2012 on the Criminal Code Chapter XVI; decree 33/2009. (X. 20.) of the Minister of Health on the clinical examination of medical devices; decree 23/2002. (V. 9.) of the Minister of Health on medical research involving humans).

### Clinical evaluation

Motor symptoms (UPDRS-III) of patients were evaluated at baseline and at the time of postoperative testing, both in off-medication state following 12-hours withdrawal of anti-parkinsonian treatment and in on-medication condition. Postoperative evaluation was performed with STN DBS stimulation turned on. Improvement in motor functions following implantation was quantified by calculating the difference between preoperative and postoperative UPDRS scores (off-medication), divided by the postoperative scores (off-medication)^75^.

### Stop Signal Reaction Time Task paradigm and behavioral analysis

The apparatus for the behavioral task consisted of 1. a custom-designed, 3D-printed ergonomic button box, featuring three buttons and containing an Arduino board (Arduino Leonardo, Arduino SRL), which detected and timestamped button presses and screen presentations; 2. a screen (size, 15.6 inches; resolution, 1366 x 768; refresh rate, 60 Hz) displaying the go signal, the stop signal and feedback; 3. a photo-sensor mounted on the upper left corner of the screen that was connected to the Arduino board to log exact times of screen presentations; 4. a notebook running custom-written, Psychtoolbox-based^76^ Matlab code that controlled running of the task; 5. a pulse train generator (Pulse Pal v2, Sanworks LLC^77^) sending TTL pulses to the data acquisition system, triggered by the output of the Arduino board, for precise synchronization of behavioral and brain data. In intraoperative recordings, the isolated TTL signal was connected to an active EMG channel of the data acquisition system; in postoperative recordings, TTL pulses were sent to a dedicated trigger channel of the EEG recording system.

We adapted the SSRT task to Parkinson’s patients by choosing the ‘go task’ to be difficult enough to obtain sufficiently long SSDs, but simple enough to be manageable for PD patients during surgery. Patients viewed visual stimuli on a screen displaying two integers between 1 and 3 presented simultaneously next to each other, and they were instructed to press the corresponding buttons in the same order as quickly as they can on a custom-designed button box (approved utility model U2200127; submitted patent application P2200356 and PCT/HU2023/050054). After patients responded by key presses, a feedback screen appeared stating whether the response was correct (over green background), or incorrect (over red background). In a subset of the trials, the screen featuring the go stimuli was followed by a ‘STOP’ signal (over red background) presented at variable delays, instructing patients to withhold button pressing. The percentage of stop trials from all presented trials within one session was 19.4 ± 7 % (mean ± SD). If patients failed to withhold button pressing, a feedback screen over red background informed them about the unsuccessful stopping. In case patients managed to refrain from pressing during a 2 second response window, a feedback screen over green background reinforced the successful stop response. The task was performed first with the right, then with the left hand preoperatively (only right hand: pd01, pd02) and also during implantation (only right hand: pd01; only left hand: pd03). Postoperative measurements started after a 12-hours withdrawal of PD medication and at least 60 minutes after DBS was turned off. Patients performed the SSRT task first with their right, then with their left hand with 60-channel EEG and EMG monitoring (only left hand: pd05, pd16; no tests: pd07). Next, the DBS stimulator was turned on and after 10 minutes of pause, the task was repeated for the right hand (pd04, pd05, pd13, pd14, pd15, pd16), the left hand (pd10) or both hands (pd01, pd02, pd08, pd12).

RT was calculated as the time interval between cue presentation and the first key press. Median RT was calculated after excluding outliers (values exceeding three scaled median absolute deviations from the median). SSDp0.5 was computed after fitting a generalized linear regression model (binary logistic regression) with response variables as successful (1) and failed stop (0) trials and corresponding SSD values as explanatory variables (Fig. 1b). SSDp0.5 corresponded to the value of 0.5 probability. Task performance was calculated as the percentage of correct trials relative to the total number of go trials (i.e., trials without a stop signal presentation). Stop performance was defined as the percentage of successful stop trials relative to the total number of stop trials.

### Neurosurgical procedure

Bilateral DBS leads (St. Jude Medical Infinity 6170 / Boston Scientific Vercise Cartesia 2202; Extended Data Table 2) and pulse generator (IPG) were implanted for each patient. For individual anatomical planning, preoperative MRI (contrast-enhanced 3D T1 weighted and T2 weighted images obtained with a 3T Phillips Achieva MRI scanner no more than six months in advance of implantation) were merged with stereotactic CT scan acquired on the day of the surgery (contrast-enhanced CT with a Leksell G frame fixed on the patient’s head). For preoperative planning, Medtronic StealthStation S7 Cranial software was employed. Standard stereotaxic principles were applied to select individual anatomical targets in the STN^78^. Trajectories were planned based on individual anatomical structures.

Dopaminergic medication was discontinued 12 h prior to the surgery. The implantation of the DBS leads was accomplished using local anesthetics, while the IPG was implanted under general anesthesia. The pre-planned trajectory was monitored with five microelectrodes (in anterior, lateral, medial, posterior and central positions), while one macroelectrode was used for delivering stimulation to assess the clinical outcome. The permanent DBS lead was implanted to the selected microelectrode trajectory. During gradual descent of the microelectrodes, the procedure was halted when the surgical team determined likely positioning in the STN with sufficient single unit quality (not necessarily at target depth).

### Microelectrode localization

Localization of the microelectrodes were reconstructed post hoc by the fusion of postoperative contrast-enhanced CT (acquired six weeks after implantation) with the preoperative T1 MRI scans, using the Medtronic StealthStation S7 software. After manually reconstructing the trajectory of the implanted electrode based on the CT artefact, the trajectories of the other 4 microelectrodes were extrapolated from the intraoperative position, the dimensions of the Ben-Gun and the depth of the final electrode with respect to the planned target position.

Additionally, diffusion tensor imaging was also performed and subregions of the STN were identified based on the connectivity profile of the STN. DICOM images were converted to compressed NIFTI format using MRICron’s dcm2niix converter. Tools available in the FMRIB Software Library (FSL 6.0.6.2) were used for linear and non-linear image registrations, and DTI data processing. Results of GPU based calculations have been obtained on a computer equipped with a Geforce Titan Xp, running Elementary OS 7. Probabilistic parcellation and anatomical classification of cortical surfaces according to the Destrieux Atlas was carried out using Freesurfer 7.3.2^79^. Classification targets for probabilistic tractography were created by combining cortical regions according to their designated functions, resulting in 3 target structures: limbic, motor, and sensory areas. The subthalamic nuclei, determined as seed regions, were manually delineated by two researchers independently on T2 images in ‘mrview’ (MRtrix 3.0.4). DTI images were corrected for motion, eddy currents and EPI distortions using tools available in FSL 6.0.6.2^80^.

B0 non-diffusion volumes were visually inspected to rule out any severe artifacts before further analysis. Estimation of diffusion tensors and crossing fibers was carried out according to methods described by Behrens et. al.^81^ using the GPU-based implementation of BedpostX with the following parameters applied: burn in value: 1000, number of fibers per voxel: 2, fudge factor: 1, applied deconvolution model: zeppelin.

Probabilistic fiber tracking was carried out using GPU-based implementation of ProbtrackX^82^ (FSL 6.0.6.2) with the following parameters applied: curvature threshold: 0.2, step length: 0.5, number of samples: 5000, volume fraction before subsidiary fiber orientations considered: 0.01, modified Euler streaming. The STN was selected as a seed region, while cortical areas described earlier were defined as classification targets for tractography in each hemisphere independently. The corpus callosum was selected as an exclusion mask to restrict fiber tracking only to the ipsilateral hemisphere. Transformation matrices between anatomical and diffusion spaces were prepared using Flirt with 6 degrees of freedom and mutual information selected as cost function. The NIFTI files containing the STN subregions (motor, limbic, associative) were loaded into Medtronic StealthStation S7 software and were assessed manually to determine whether the microelectrode measurement points reached each subregion. The center of gravities (CoGs, centroids) of the STN subregions were calculated with fslmaths (FSL 6.0.6.2) and 3D Euclidean distances were determined between the microelectrode contact coordinates and the CoGs of the STN subregions.

Limitations of microeletrode reconstruction. Microelectrode reconstructions were based on the standard intraoperative positioning of the Ben-Gun; however, we lacked intraoperative imaging to confirm the reconstructed trajectories. This limitation may introduce inaccuracies in the calculated 3D distances between the CoG of the STN subregions and the recording sites. Additionally, localization of postoperatively stimulated contacts was calculated as the inner center of the full ring; therefore, single segments of directional DBS electrodes could not be differentiated.

### Electrophysiological data acquisition and preprocessing of LFP/EEG data

Intraoperative microelectrode recordings were performed with the clinical microelectrode recording system (ISIS Microelectrode Recording (MER) System, Innomed Medizintechnik GmbH, Emmendingen, Germany), with 20 kHz sampling frequency. Additionally, two EEG electrodes placed at positions corresponding to F3 and F4 of the 10-20 system were also recording brain activity (connected to the EMG headbox module of the same MER system), resulting in one bipolar EEG derivation (F4-F3).

Postoperative EEG was recorded with a standard 60-channel recording system and EEG cap (Brain Products Co., Munich, Germany), with 2048 Hz sampling rate. Electrodes were positioned according to the extended 10–20 system.

As a first step, electrophysiological data was synchronized with behavioral events, based on timestamps saved by the electrophysiological recording system and by the experimental setup, respectively. Time series were aligned, and behavioral timestamps were converted to the MER/EEG system’s clock. Logged events (go and stop stimuli) were matched with the corresponding MER/EEG trigger using a 50 ms tolerance. This resulted in a few unmatched events due to broken or undetected TTL pulses, for which timestamps were linearly interpolated using the matched timestamps.

In case of postoperative recordings obtained during DBS stimulation, the stimulation artefacts were removed using DBSfilt, an open-source Matlab toolbox developed for this purpose^83^. All data were re-sampled to 250 Hz and filtered using a 100 Hz low-pass filter (‘sinc’ FIR filter using a Hamming window with default parameters of the pop_eegfiltnew.m Matlab function of the EEGLab package^84^). Line noise was removed either by CleanLine filter^85^ or by 45-55 Hz notch filter, if irregular noise escaped the CleanLine filter. Next, the data was epoched according to the synchronized behavioral events to time windows of −2 to 2 s around each behaviorally relevant event (go signal, first key press, stop signal, feedback). Epoched data was revised visually and trials with high amplitude noise were rejected manually.

In case of postoperative scalp EEG recordings, artifact-rejected data were high-pass filtered above 2 Hz for Independent Component Analyses (ICA)^86^, by implementing fastICA algorithm^87^ (EEGLab plugin). ICA components associated with eye movements, faulty electrodes, stereotypic muscle artefacts and/or pulse artefacts were rejected. Rejected ICA components were removed from data filtered above 0.5 Hz, to regain low-frequency components^86^. Average baseline (−2 to −1 s) amplitude was subtracted from each data epoch. Following these steps, scalp current densities^88,89^ were calculated by using FieldTrip Matlab toolbox^90^ (ft_scalpcurrentdensity)^91^, implementing the spherical spine method^92^.

### Data analysis of LFP/ EEG data

All preprocessing and data analyses were performed using open-source and custom-written Matlab algorithms (MATLAB Version: 9.7.0 (R2019b), The MathWorks Inc.)^93^. Time-frequency decompositions were performed on concatenated and z-scored data epochs using Morlet wavelets (cycle number, 6^94^), employing the algorithm of Torrence and Compo^95^. Individual time-frequency epochs were normalized for each frequency component using the full length of the epoch^96^. Event-related spectrograms were visualized as power changes relative to baseline (−1 to −0.5 s). When comparing conditions, a common baseline across the compared conditions was applied.

When analyzing STN LFP, time-frequency measures were averaged across all microelectrode channels. The dominant frequency range within the delta band (1-4 Hz) was determined around go and stop signals (−2 to 2 s windows) separately for calculating spike-phase coupling. The frequency component with maximal power (Frpeak) after averaging across time and epochs was determined. Dominant frequency range was defined as Frpeak ± Frpeak/6.

Magnitude-squared wavelet coherence (MSWC) was calculated between each microelectrode channel and the F4-F3 derivation using the built-in Matlab function wcoherence.m. Similarly to spectrograms, MSWC maps were also averaged across microelectrode channels.

### Single- and multiunit activity analysis

Clustering of action potentials (Fig. 4b-e) was performed off-line on the 20 kHz sampled MER recordings, using the MClust 3.5 Matlab-based spike sorting toolbox (A.D. Redish, https://github.com/adredish/MClust-Spike-Sorting-Toolbox.git)^97^. First, the automated KlustaKwik^98^ algorithm was run to create 2 to 7 clusters using amplitude (peak-to-valley), waveform energy and the first three principal component of the waveform (‘WavePC1-3’) as features. Next, clusters were manually inspected and (i) accepted (sufficient waveform quality and low proportion of refractory period violations in the autocorrelograms), (ii) rejected (insufficient waveform quality due to low amplitude, high variance or strong drift, or high proportion of refractory period violations in the autocorrelograms), (iii) merged (if the cross-correlogram and temporal evolution confirmed the identity of the units) or (iv) further split by running KlustaKwik on the selected cluster (if multiple distinct waveforms could be observed within the cluster). Waveform distributions of the resulting accepted clusters were inspected, and clearly identifiable cluster contaminations were removed. Isolation distance and L-ratio was calculated using WavePC1-3 and waveform energy as features, as suggested by Schmitzer-Torbert et al.^99^ for comparability across studies^15,68,100,101^. Clusters with Isolation Distance > 15 and L-ratio < 0.02 were considered single units; all other accepted clusters were considered multiunits.

Clustered units were examined in terms of their responsiveness to behavioral events, using the CellBase open-source Matlab toolbox^102^ (https://github.com/hangyabalazs/CellBase.git). The action potential times were aligned to task events (go signal, key press, stop signal and feedback), resulting in peri-event time histograms (PETH). The activity in the baseline window (−2.5 to −1 s relative to go signal; −3 to −1.5 s relative to key press and stop; 1.5 to 3 s relative to feedback) was compared to a test window (0 to 1 s for all events). First, local extremes were found and checked whether they surpassed (possible activation) or fell below (possible inhibition) mean baseline spiking probability. Then, a smaller window around the local extreme was defined, as the interval between the timepoints where the PETH crossed half-distances between the local extreme and baseline average, respectively. The spike count in the resulting activation/inhibition time interval was compared to a matching baseline time interval, selected with an identical method within the predefined baseline window, using Mann-Whitney U test (one-sided, due to the asymmetric alternative hypothesis). Spiking activity corresponding to stop trials of different outcome was assessed by calculating PETHs derived from trials partitioned to failed and successful stop trials.

Coupling of spiking activity to LFP delta phase was also investigated. Spikes within the interval of −1.5 to 1.5 s around the go or the stop signal were selected and pooled from all respective epochs. LFP recorded on the same channel as the examined unit was band-pass filtered within the dominant delta frequency band (see above) and then Hilbert-transformed. The phase angles of the resulting analytic signal were extracted. Phase angles at timepoints closest to spike times were collected. The angle and the magnitude of the first trigonometric moment of this phase distribution defined the preferred phase and coupling strength (mean resultant vector length, MRL) of the examined unit, respectively. Preferred phases of units significantly coupled to LFP delta (Rayleigh’s test for circular uniformity, p < 0.05) were pooled to form a population phase histogram. MRLs of these units characterized the degree of LFP delta coupling at the population level.

Burstiness was estimated by the presence of a short-lag peak (0-10 ms) in the autocorrelograms (AC) of the units. It was quantified by a bursting index, calculated by subtracting the AC counts in a ‘baseline’ latency window (876-1125 ms, chosen as to eliminate potential dependence on delta-rhythmic fluctuations of spiking) from the counts in the 0-10 ms period and dividing the difference with the larger of the two counts. Units were grouped into ‘bursting’ and ‘non-bursting’ groups based on an empirical threshold of 0.35^103–105^. Additionally, ‘strongly bursting’ units were compared to ‘weakly bursting’ units by a median split of the burst indices.

### Statistics

Behavioral parameters were compared with Kruskal-Wallis and post-hoc Tukey-Kramer tests. For comparison within patient groups, Mann-Whitney U test was applied.

Correlations of various task-, clinical- and electrophysiological parameters were quantified using Pearson’s correlation and tested with t-test. In case of correlating delta power with RT, time-frequency correlation maps were obtained (correlation was calculated for each individual time-frequency datapoint within the selected epoch).

Changes relative to baseline for time-frequency power coefficients, and wavelet coherence maps were tested using permutation statistics (n = 1000 iterations, p < 0.05) with false discovery rate correction. For time-frequency maps associated with successful / failed stop trials, permutation statistics (n = 1000 iterations) with cluster-based correction was applied (p < 0.05), using Fieldtrip toolbox. Similarly, PETHs of successful/failed stop trials and low/high burst index units were also compared by permutation statistics with the same parameters. Difference between average delta power of successful and failed stop trials (in selected time window and frequency range) was tested with Mann-Whitney U-test (p < 0.05).

For comparisons of number of units in various STN subregions, Fisher’s exact test was applied. Coupling of spikes to LFP delta was considered significant if the null hypothesis of uniform distribution of phase angles was rejected by Rayleigh’s test for circular uniformity (p < 0.05). Circular distributions of preferred phases were compared with Watson’s two sample test of homogeneity (p < 0.05).

## Data availability

Deidentified human electrophysiology data will be made available upon publication.

## Code availability

Unique code developed to analyze these data is available at https://github.com/hangyabalazs/human-STN-delta.

## Acknowledgement

We thank prof. Dániel Bereczki for his support of the project. This work was supported by the Hungarian Brain Research Program NAP3.0 (NAP2022-I-1/2022) of the Hungarian Academy of Sciences and the ERC POC grant 101123104 to B.H.

## References

1. Jahfari, S. et al. Effective Connectivity Reveals Important Roles for Both the Hyperdirect (Fronto-Subthalamic) and the Indirect (Fronto-Striatal-Pallidal) Fronto-Basal Ganglia Pathways during Response Inhibition. The Journal of Neuroscience 31, 6891–6899 (2011).

2. Schmidt, R., Leventhal, D. K., Mallet, N., Chen, F. & Berke, J. D. Canceling actions involves a race between basal ganglia pathways. Nat Neurosci 16, 1118–1124 (2013).

3. Bari, A. & Robbins, T. W. Inhibition and impulsivity: behavioral and neural basis of response control. Prog Neurobiol 108, 44–79 (2013).

4. Herz, D. M. et al. Motivational Tuning of Fronto-Subthalamic Connectivity Facilitates Control of Action Impulses. The Journal of Neuroscience 34, 3210–3217 (2014).

5. Perugini, A., Ditterich, J. & Basso, M. A. Patients with Parkinson’s Disease Show Impaired Use of Priors in Conditions of Sensory Uncertainty. Current Biology 26, 1902–1910 (2016).

6. Zhang, Q. et al. Low-frequency oscillations link frontal and parietal cortex with subthalamic nucleus in conflicts. Neuroimage 258, 119389 (2022).

7. Chang, A., Ide, J. S., Li, H.-H., Chen, C.-C. & Li, C.-S. R. Proactive Control: Neural Oscillatory Correlates of Conflict Anticipation and Response Slowing. eNeuro 4, ENEURO.0061-17.2017 (2017).

8. Nguyen, T. Van, Balachandran, P., Muggleton, N. G., Liang, W.-K. & Juan, C.-H. Dynamical EEG Indices of Progressive Motor Inhibition and Error-Monitoring. Brain Sci 11, 478 (2021).

9. Khan, A. U. et al. Low-Frequency Oscillations in Mid-rostral Dorsolateral Prefrontal Cortex Support Response Inhibition. The Journal of Neuroscience 44, e0122242024 (2024).

10. Ghahremani, A. et al. Event-related deep brain stimulation of the subthalamic nucleus affects conflict processing. Ann Neurol 84, 515–526 (2018).

11. Zavala, B. A. et al. Midline Frontal Cortex Low-Frequency Activity Drives Subthalamic Nucleus Oscillations during Conflict. The Journal of Neuroscience 34, 7322–7333 (2014).

12. Cavanagh, J. F. et al. Subthalamic nucleus stimulation reverses mediofrontal influence over decision threshold. Nat Neurosci 14, 1462–1467 (2011).

13. Chen, W. et al. Prefrontal-Subthalamic Hyperdirect Pathway Modulates Movement Inhibition in Humans. Neuron 106, 579–588.e3 (2020).

14. Mosher, C. P., Mamelak, A. N., Malekmohammadi, M., Pouratian, N. & Rutishauser, U. Distinct roles of dorsal and ventral subthalamic neurons in action selection and cancellation. Neuron 109, 869–881.e6 (2021).

15. London, D. et al. Distinct population code for movement kinematics and changes of ongoing movements in human subthalamic nucleus. Elife 10, (2021).

16. Rogers, K., Gold, J. & Ding, L. The Subthalamic Nucleus Contributes Causally to Perceptual Decision-Making in Monkeys. bioRxiv: the preprint server for biology (2024). doi:10.1101/2024.04.09.588715.

17. Lourens, M. A. J. et al. Functional neuronal activity and connectivity within the subthalamic nucleus in Parkinson’s disease. Clinical Neurophysiology 124, 967–981 (2013).

18. Steigerwald, F. et al. Neuronal Activity of the Human Subthalamic Nucleus in the Parkinsonian and Nonparkinsonian State. J Neurophysiol 100, 2515–2524 (2008).

19. Pagnier, G. J., Asaad, W. F. & Frank, M. J. Double dissociation of dopamine and subthalamic nucleus stimulation on effortful cost/benefit decision making. Current Biology 34, 655–660.e3 (2024).

20. Herz, D. M. et al. Mechanisms Underlying Decision-Making as Revealed by Deep-Brain Stimulation in Patients with Parkinson’s Disease. Current Biology 28, 1169–1178.e6 (2018).

21. Krack, P., et al. Five-Year Follow-up of Bilateral Stimulation of the Subthalamic Nucleus in Advanced Parkinson’s Disease From the Departments of Clinical and Bio-Logical Neurosciences (P. N Engl J Med vol. 349 www.nejm.org (2003).

22. Herz, D. M. Neuroscience: Therapy modulates decision-making in Parkinson’s disease. Current Biology 34, R148–R150 (2024).

23. Verbruggen, F. & Logan, G. D. Automatic and Controlled Response Inhibition: Associative Learning in the Go/No-Go and Stop-Signal Paradigms. J Exp Psychol Gen 137, 649–672 (2008).

24. Duque, J., Olivier, E. & Rushworth, M. Top–Down Inhibitory Control Exerted by the Medial Frontal Cortex during Action Selection under Conflict. J Cogn Neurosci 25, 1634–1648 (2013).

25. Chen, W. et al. Prefrontal-Subthalamic Hyperdirect Pathway Modulates Movement Inhibition in Humans. Neuron 106, 579–588 e3 (2020).

26. Senzai, Y., Fernandez-Ruiz, A. & Buzsáki, G. Layer-Specific Physiological Features and Interlaminar Interactions in the Primary Visual Cortex of the Mouse. Neuron 101, 500–513.e5 (2019).

27. Ammari, R., Bioulac, B., Garcia, L. & Hammond, C. The Subthalamic Nucleus becomes a Generator of Bursts in the Dopamine-Depleted State. Its High Frequency Stimulation Dramatically Weakens Transmission to the Globus Pallidus. Front Syst Neurosci 5, 43 (2011).

28. Tai, C. H. Subthalamic burst firing: A pathophysiological target in Parkinson’s disease. Neuroscience and Biobehavioral Reviews vol. 132 410–419 Preprint at 10.1016/j.neubiorev.2021.11.044 (2022).

29. Royer, S. et al. Control of timing, rate and bursts of hippocampal place cells by dendritic and somatic inhibition. Nat Neurosci 15, 769–775 (2012).

30. Hegedüs, P., Heckenast, J. & Hangya, B. Differential recruitment of ventral pallidal e-types by behaviorally salient stimuli during Pavlovian conditioning. iScience 24, (2021).

31. Laszlovszky, T. et al. Distinct synchronization, cortical coupling and behavioral function of two basal forebrain cholinergic neuron types. Nat Neurosci 23, 992–1003 (2020).

32. Kaku, H. et al. Unsupervised clustering reveals spatially varying single neuronal firing patterns in the subthalamic nucleus of patients with Parkinson’s disease. Clin Park Relat Disord 3, 100032 (2020).

33. Frank, M. J., Samanta, J., Moustafa, A. A. & Sherman, S. J. Hold Your Horses: Impulsivity, Deep Brain Stimulation, and Medication in Parkinsonism. Science (1979) 318, 1309–1312 (2007).

34. Swann, N. et al. Deep brain stimulation of the subthalamic nucleus alters the cortical profile of response inhibition in the beta frequency band: A scalp EEG study in parkinson’s disease. Journal of Neuroscience 31, 5721–5729 (2011).

35. Mirabella, G. et al. Deep brain stimulation of subthalamic nuclei affects arm response inhibition in Parkinson’s patients. Cereb Cortex 22, 1124–1132 (2012).

36. Ray, N. J. et al. The role of the subthalamic nucleus in response inhibition: Evidence from deep brain stimulation for Parkinson’s disease. Neuropsychologia 47, 2828–2834 (2009).

37. van den Wildenberg, W. P. M., et al. Deep-brain stimulation of the subthalamic nucleus improves overriding motor actions in Parkinson’s disease. Behavioural Brain Research 402, 113124 (2021).

38. Obeso, I., Wilkinson, L., Rodríguez-Oroz, M. C., Obeso, J. A. & Jahanshahi, M. Bilateral stimulation of the subthalamic nucleus has differential effects on reactive and proactive inhibition and conflict-induced slowing in Parkinson’s disease. Exp Brain Res 226, 451–462 (2013).

39. van den Wildenberg, W. P. M., et al. Stimulation of the Subthalamic Region Facilitates the Selection and Inhibition of Motor Responses in Parkinson’s Disease. J Cogn Neurosci 18, 626–636 (2006).

40. Ballanger, B. et al. Stimulation of the subthalamic nucleus and impulsivity: Release your horses. Ann Neurol 66, 817–824 (2009).

41. Eagle, D. M. et al. Stop-signal reaction-time task performance: Role of prefrontal cortex and subthalamic nucleus. Cerebral Cortex 18, 178–188 (2008).

42. Mirabella, G. et al. Stimulation of Subthalamic Nuclei Restores a Near Normal Planning Strategy in Parkinson’s Patients. PLoS One 8, e62793-(2013).

43. Wylie, S. A. et al. Subthalamic nucleus stimulation influences expression and suppression of impulsive behaviour in Parkinson’s disease. Brain 133, 3611–3624 (2010).

44. Mancini, C. et al. Unilateral Stimulation of Subthalamic Nucleus Does Not Affect Inhibitory Control. Front Neurol 9, (2019).

45. Hallett, M. Clinical neurophysiology of akinesia. Rev Neurol (Paris) 146 10, 585–90 (1990).

46. Baunez, C. & Robbins, T. W. Bilateral Lesions of the Subthalamic Nucleus Induce Multiple Deficits in an Attentional Task in Rats. European Journal of Neuroscience vol. 9 (1997).

47. Uslaner, J. M. & Robinson, T. E. Subthalamic nucleus lesions increase impulsive action and decrease impulsive choice - Mediation by enhanced incentive motivation? European Journal of Neuroscience 24, 2345–2354 (2006).

48. Serranová, T. et al. Subthalamic nucleus stimulation affects incentive salience attribution in Parkinson’s disease. Movement Disorders 26, 2260–2266 (2011).

49. Cavanagh, J. F. et al. Subthalamic nucleus stimulation reverses mediofrontal influence over decision threshold. Nat Neurosci 14, 1462–1467 (2011).

50. Chang, A., Ide, J. S., Li, H. H., Chen, C. C. & Li, C. R. Proactive Control: Neural Oscillatory Correlates of Conflict Anticipation and Response Slowing. eNeuro 4, (2017).

51. Cohen, M. X. & van Gaal, S. Subthreshold muscle twitches dissociate oscillatory neural signatures of conflicts from errors. Neuroimage 86, 503–513 (2014).

52. Lavallee, C. F., Meemken, M. T., Herrmann, C. S. & Huster, R. J. When holding your horses meets the deer in the headlights: time-frequency characteristics of global and selective stopping under conditions of proactive and reactive control. Front Hum Neurosci 8, 994 (2014).

53. Zhang, Q. et al. Low-frequency oscillations link frontal and parietal cortex with subthalamic nucleus in conflicts. Neuroimage 258, 119389 (2022).

54. Green, N. et al. Reduction of influence of task difficulty on perceptual decision making by stn deep brain stimulation. Current Biology 23, 1681–1684 (2013).

55. Nambu, A., Tokuno, H. & Takada, M. Functional significance of the cortico–subthalamo–pallidal ‘hyperdirect’ pathway. Neurosci Res 43, 111–117 (2002).

56. Tan, H. et al. Decoding gripping force based on local field potentials recorded from subthalamic nucleus in humans. Elife 5, 1–24 (2016).

57. Hell, F., Taylor, P. C. J., Mehrkens, J. H. & Bötzel, K. Subthalamic stimulation, oscillatory activity and connectivity reveal functional role of STN and network mechanisms during decision making under conflict. Neuroimage 171, 222–233 (2018).

58. Frank, M. J. Hold your horses: A dynamic computational role for the subthalamic nucleus in decision making. Neural Networks 19, 1120–1136 (2006).

59. Pasquereau, B. & Turner, R. S. Neural dynamics underlying self-control in the primate subthalamic nucleus. Elife 12, 1–27 (2023).

60. Patel, S. R. et al. Studying task-related activity of individual neurons in the human brain. Nat Protoc 8, 949–957 (2013).

61. Rossi, P. J. et al. The human subthalamic nucleus and globus pallidus internus differentially encode reward during action control. Hum Brain Mapp 38, 1952–1964 (2017).

62. Bastin, J. et al. Inhibitory control and error monitoring by human subthalamic neurons. Transl Psychiatry 4, (2014).

63. Zaghloul, K. A. et al. Neuronal Activity in the Human Subthalamic Nucleus Encodes Decision Conflict during Action Selection. The Journal of Neuroscience 32, 2453–2460 (2012).

64. Zavala, B. et al. Human subthalamic nucleus theta and beta oscillations entrain neuronal firing during sensorimotor conflict. Cerebral Cortex 27, 496–508 (2015).

65. Burbaud, P. et al. Neuronal activity correlated with checking behaviour in the subthalamic nucleus of patients with obsessive-compulsive disorder. Brain 136, 304–317 (2013).

66. Favre, E., Ballanger, B., Thobois, S., Broussolle, E. & Boulinguez, P. Deep Brain Stimulation of the Subthalamic Nucleus, but not Dopaminergic Medication, Improves Proactive Inhibitory Control of Movement Initiation in Parkinson’s Disease. Neurotherapeutics 10, 154–167 (2013).

67. Pasquereau, B. & Turner, R. S. A selective role for ventromedial subthalamic nucleus in inhibitory control. Elife 6, 1–27 (2017).

68. Lipski, W. J. et al. Subthalamic nucleus neurons differentially encode early and late aspects of speech production. Journal of Neuroscience 38, 5620–5631 (2018).

69. Lévesque, J.-C. & Parent, A. GABAergic interneurons in human subthalamic nucleus. Movement Disorders 20, 574–584 (2005).

70. Micheli, F. et al. Impulsivity Markers in Parkinsonian Subthalamic Single-Unit Activity. Movement Disorders 36, 1435–1440 (2021).

71. Hershey, T. et al. Mapping Go-No-Go performance within the subthalamic nucleus region. Brain 133, 3625–3634 (2010).

72. van Wouwe, N. C. et al. Focused stimulation of dorsal subthalamic nucleus improves reactive inhibitory control of action impulses. Neuropsychologia 99, 37–47 (2017).

73. Schuepbach, W. M. M. et al. Neurostimulation for Parkinson’s Disease with Early Motor Complications. New England Journal of Medicine 368, 610–622 (2024).

74. Hacker, M. L. et al. Deep brain stimulation in early-stage Parkinson disease. Neurology 95, e393– e401 (2020).

75. Qi, R. et al. Outcomes of STN-DBS in PD Patients With Different Rates of Disease Progression Over One Year of Follow-Up. Front Neurol 11, (2020).

76. Kleiner, M. et al. What’s new in Psychtoolbox-3. Perception 36, 1–16 (2007).

77. Sanders, J. & Kepecs, A. A low-cost programmable pulse generator for physiology and behavior. Front Neuroeng 7, 43 (2014).

78. Andrade-Souza, Y. M. et al. COMPARISON OF THREE METHODS OF TARGETING THE SUBTHALAMIC NUCLEUS FOR CHRONIC STIMULATION IN PARKINSON’S DISEASE. Neurosurgery 62, (2008).

79. Fischl, B. FreeSurfer. NeuroImage vol. 62 774–781 Preprint at 10.1016/j.neuroimage.2012.01.021 (2012).

80. Jenkinson, M., Beckmann, C. F., Behrens, T. E. J., Woolrich, M. W. & Smith, S. M. FSL. Neuroimage 62, 782–790 (2012).

81. Behrens, T. E. J. et al. Non-invasive mapping of connections between human thalamus and cortex using diffusion imaging. Nat Neurosci 6, 750–757 (2003).

82. Hernandez-Fernandez, M. et al. Using GPUs to accelerate computational diffusion MRI: From microstructure estimation to tractography and connectomes. Neuroimage 188, 598–615 (2019).

83. Lio, G., Thobois, S., Ballanger, B., Lau, B. & Boulinguez, P. Removing deep brain stimulation artifacts from the electroencephalogram: Issues, recommendations and an open-source toolbox. Clin Neurophysiol 129, 2170–2185 (2018).

84. Delorme, A. & Makeig, S. EEGLAB: An Open Source Toolbox for Analysis of Single-Trial EEG Dynamics Including Independent Component Analysis. Journal of Neuroscience Methods vol. 134 http://www.sccn.ucsd.edu/eeglab/ (2004).

85. Mullen, T. CleanLine EEGLAB plugin. San Diego, CA: Neuroimaging Informatics Toolsand Resources Clearinghouse (NITRC) (2012).

86. Winkler, I., Debener, S., Müller, K.-R. & Tangermann, M. On the Influence of High-Pass Filtering on ICA-Based Artifact Reduction in EEG-ERP. (2015). doi:10.0/Linux-x86_64.

87. Hyvärinen, A. Fast and Robust Fixed-Point Algorithms for Independent Component Analysis. IEEE TRANSACTIONS ON NEURAL NETWORKS vol. 10 (1999).

88. Nunez, P. L. & Pilgreen, K. L. The Spline-Laplacian in Clinical Neurophysiology: A Method to Improve EEG Spatial Resolution. Journal of Clinical Neurophysiology 8, (1991).

89. Perrin, F., Bertrand, O. & Pernier, J. Scalp Current Density Mapping: Value and Estimation from Potential Data. IEEE Trans Biomed Eng BME-34, 283–288 (1987).

90. Oostenveld, R., Fries, P., Maris, E. & Schoffelen, J. M. FieldTrip: Open source software for advanced analysis of MEG, EEG, and invasive electrophysiological data. Comput Intell Neurosci 2011, (2011).

91. Waschke, L. et al. Modality-specific tracking of attention and sensory statistics in the human electrophysiological spectral exponent. doi:10.7554/eLife.

92. Perrin, F., Pernier, J., Bertrand, O. & Echallier, J. F. Spherical splines for scalp potential and current density mapping. Electroencephalogr Clin Neurophysiol 72, 184–187 (1989).

93. Inc., T. M. MATLAB version: 9.7.0 (R2019b). Preprint at https://www.mathworks.com (2019).

94. Farge, M. Wavelet Transforms and their Applications to Turbulence. Annu Rev Fluid Mech 24, 395–457 (1992).

95. Torrence, C. & Compo, G. P. A Practical Guide to Wavelet Analysis. Bull Am Meteorol Soc 79, 61– 78 (1998).

96. Grandchamp, R. & Delorme, A. Single-trial normalization for event-related spectral decomposition reduces sensitivity to noisy trials. Front Psychol 2, 236 (2011).

97. Hangya, B., Ranade, S. P., Lorenc, M. & Kepecs, A. Central Cholinergic Neurons Are Rapidly Recruited by Reinforcement Feedback. Cell 162, 1155–1168 (2015).

98. Rossant, C. et al. Spike sorting for large, dense electrode arrays. Nat Neurosci 19, 634–641 (2016).

99. Schmitzer-Torbert, N., Jackson, J., Henze, D., Harris, K. & Redish, A. D. Quantitative measures of cluster quality for use in extracellular recordings. Neuroscience 131, 1–11 (2005).

100. Kim, J. et al. Inhibitory Basal Ganglia Inputs Induce Excitatory Motor Signals in the Thalamus. Neuron 95, 1181–1196.e8 (2017).

101. Tekriwal, A., Lintz, M. J., Thompson, J. A. & Felsen, G. Disrupted basal ganglia output during movement preparation in hemiparkinsonian mice is consistent with behavioral deficits. J Neurophysiol 126, 1248–1264 (2021).

102. Hegedüs, P., Sviatkó, K., Király, B., Martínez-Bellver, S. & Hangya, B. Cholinergic activity reflects reward expectations and predicts behavioral responses. iScience 26, 105814 (2023).

103. Laszlovszky, T. et al. Distinct synchronization, cortical coupling and behavioral function of two basal forebrain cholinergic neuron types. Nat Neurosci 23, 992–1003 (2020).

104. Hegedüs, P., Heckenast, J. & Hangya, B. Differential recruitment of ventral pallidal e-types by behaviorally salient stimuli during Pavlovian conditioning. iScience 24, 102377 (2021).

105. Hegedüs, P. et al. Parvalbumin-expressing basal forebrain neurons mediate learning from negative experience. Nat Commun 15, 4768 (2024).

