## Supplementary Figures and Tables for "Neurons of the human subthalamic nucleus engage with local delta frequency processes during action cancellation"

### Supplementary Material

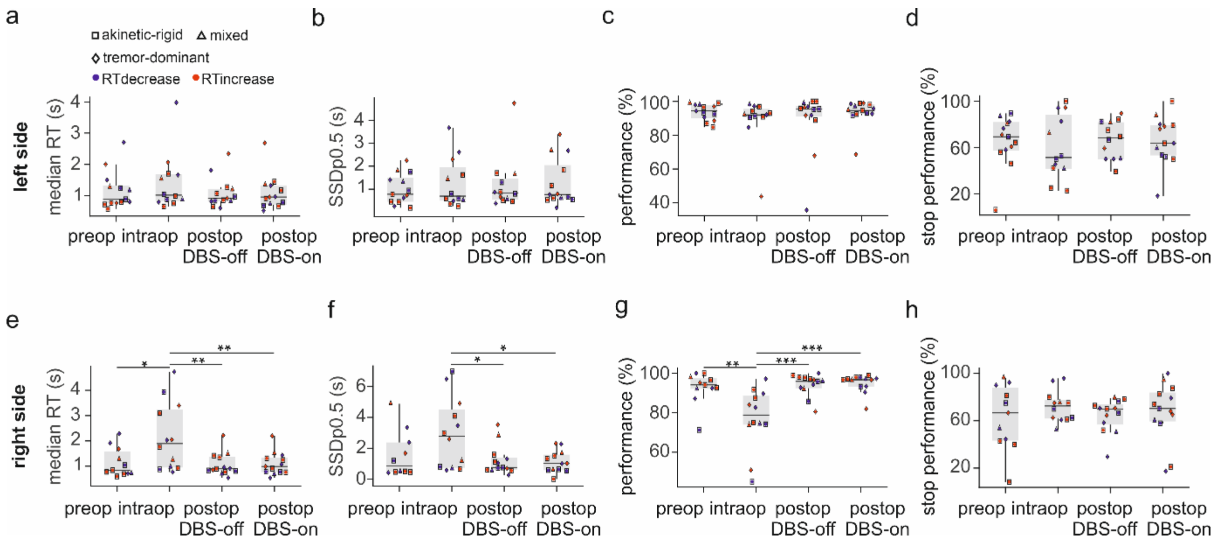


**Extended Data Figure 1. Behavioral data of patients with PD performing the Stop Signal Reaction Time task. a)** Patient reaction times in preoperative, intraoperative, and postoperative left side tests. Blue, patients with postoperative RT increase; red, patients with RT decrease; diamond, patients with tremor-dominant PD; square, patients with akinetic-rigid PD; triangle, patients with mixed PD. Box-whisker plots show median, interquartile range and non-outlier range. **b)** SSDp0.5 distributions across patients in preoperative, intraoperative, and postoperative left side tests. **c)** Task performance distributions across patients in preoperative, intraoperative, and postoperative left side tests. **d)** Stop performance distributions across patients in preoperative, intraoperative, and postoperative left side tests. **e-h)** The same as a-d, but for right side tests. Intraoperative RT and SSDp0.5 were significantly higher, and task performance was significantly lower than pre- and postoperative values in case of right side tests (Kruskal-Wallis test with post-hoc Tukey-Kramer test; *, $p < 0.05$; **, $p < 0.01$; ***, $p < 0.001$).

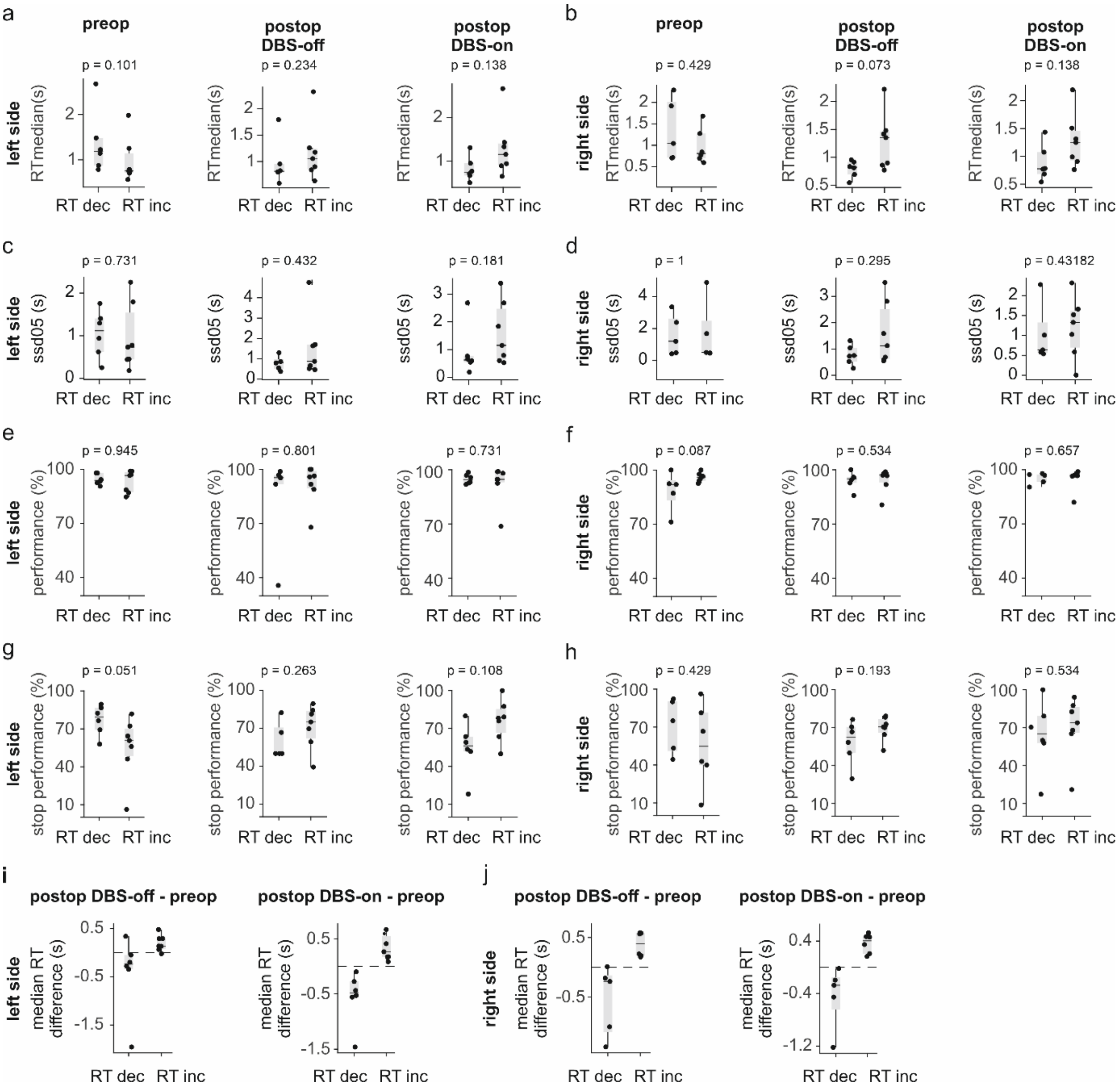


**Extended Data Figure 2. Comparison of SSRT parameters based on the sign of pre- to postoperative RT change. a)** Median RT values of patients with RT decrease (dec.) and RT increase (inc.) in preoperative (left), postoperative DBS-off (middle) and DBS-on (right) left side measurements. Box-whisker plots show median, interquartile range and non-outlier range. **b)** The same as in panel a, but for right side measurements. **c)** SSDp0.5 values of patients with RT decrease and RT increase in preoperative (left), postoperative DBS-off (middle) and DBS-on (right) left side measurements. **d)** The same as in panel c, but for right side measurements. **e)** Task performance of patients with RT decrease and RT increase in preoperative (left), postoperative DBS-off (middle) and DBS-on (right) left side measurements. **f)** The same as in panel e, but for right side measurements. **g)** Stop performance of patients with RT decrease and RT increase in preoperative (left), postoperative DBS-off (middle) and DBS-on (right) left side measurements. **h)** The same as in panel g, but for right side measurements. **i)** Magnitude of pre- to postoperative RT change of patients with RT decrease and RT increase, calculated based on postoperative DBS-off (left) or DBS-on (right) left side measurements. **j)** The same as in panel i, but for right side measurements.

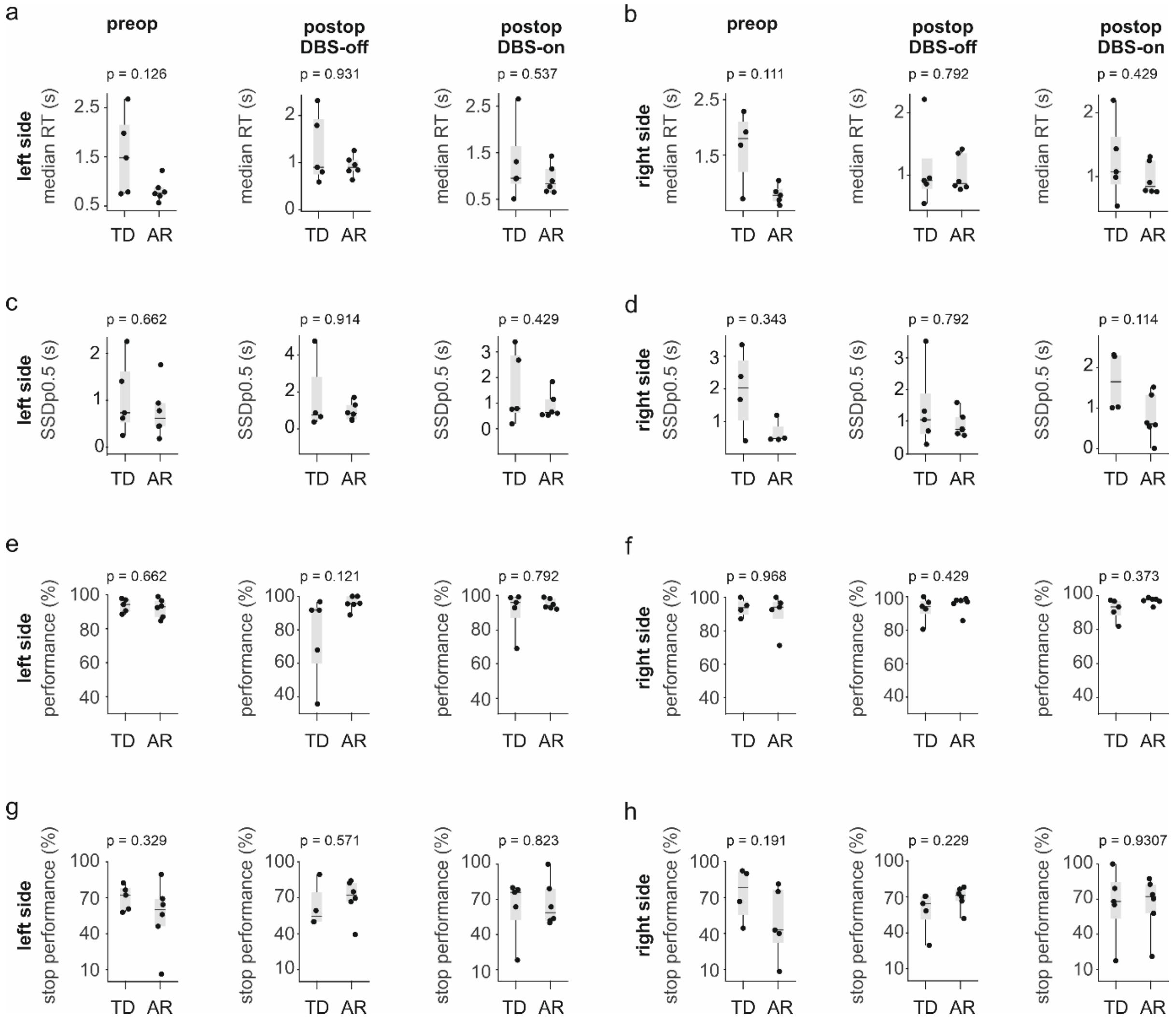


**Extended Data Figure 3. Comparison of behavioral parameters of patients with different clinical phenotypes. a)** Median RT values of patients with tremor-dominant (TD) and akinetic-rigid (AR) PD in preoperative (left), postoperative DBS-off (middle) and DBS-on (right) left side measurements. Box-whisker plots show median, interquartile range and non-outlier range. **b)** The same as in panel a, but for right side measurements. **c)** SSDp0.5 values of patients with tremor-dominant and akinetic-rigid PD in preoperative (left), postoperative DBS-off (middle) and DBS-on (right) left side measurements. **d)** The same as in panel c, but for right side measurements. **e)** Task performance of patients with tremor-dominant and akinetic-rigid PD in preoperative (left), postoperative DBS-off (middle) and DBS-on (right) left side measurements. **f)** The same as in panel e, but for right side measurements. **g)** Stop performance of patients with tremor-dominant and akinetic-rigid PD in preoperative (left), postoperative DBS-off (middle) and DBS-on (right) left side measurements. **h)** The same as in panel g, but for right side measurements. Although no significant differences were found in any of the presented behavioral parameters (Mann-Whitney U-test), we note that patients with akinetic-rigid PD showed less variable RTs.

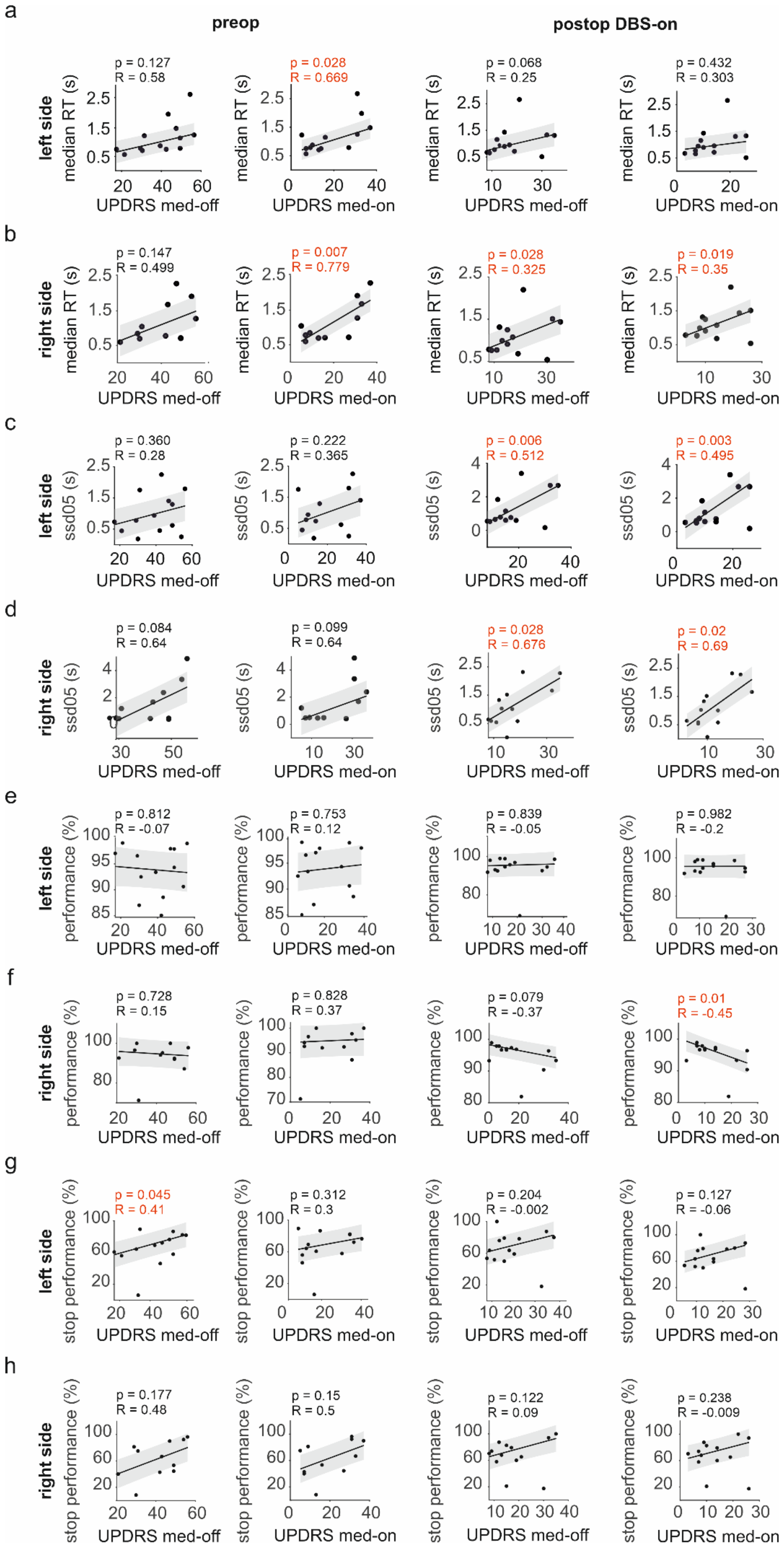
a
preop
postop DBS-on
left side
median RT (s)
p = 0.127
R = 0.58
p = 0.028
R = 0.669
p = 0.068
R = 0.25
p = 0.432
R = 0.303
UPDRS med-off
UPDRS med-on
b
right side
p = 0.147
R = 0.499
p = 0.007
R = 0.779
p = 0.028
R = 0.325
p = 0.019
R = 0.35
c
ssd05 (s)
p = 0.360
R = 0.28
p = 0.222
R = 0.365
p = 0.006
R = 0.512
p = 0.003
R = 0.495
d
p = 0.084
R = 0.64
p = 0.099
R = 0.64
p = 0.028
R = 0.676
p = 0.02
R = 0.69
e
performance (%)
p = 0.812
R = -0.07
p = 0.753
R = 0.12
p = 0.839
R = -0.05
p = 0.982
R = -0.2
f
p = 0.728
R = 0.15
p = 0.828
R = 0.37
p = 0.079
R = -0.37
p = 0.01
R = -0.45
g
stop performance (%)
p = 0.045
R = 0.41
p = 0.312
R = 0.3
p = 0.204
R = -0.002
p = 0.127
R = -0.06
h
p = 0.177
R = 0.48
p = 0.15
R = 0.5
p = 0.122
R = 0.09
p = 0.238
R = -0.009

**Extended Data Figure 4. Correlation of behavioral parameters with UPDRS scores. a)** Median RT values as a function of UPDRS scores without and with pharmacotherapy (med-off and med-on), in preoperative (left) and postoperative DBS-on (right) left side measurements. Lines indicate robust regression with 95% confidence intervals (grey shading). Pearson's correlation coefficients and corresponding p-values (t-test) are shown for each plot. **b)** The same as in panel a, but for right side measurements. **c)** SSDp0.5 values as a function of UPDRS scores without and with pharmacotherapy (med-off and med-on), in preoperative (left) and postoperative DBS-on (right) left side measurements**. d)** The same as in panel c, but for right side measurements. **e)** Task performance as a function of UPDRS scores without and with pharmacotherapy (med-off and med-on), in preoperative (left) and postoperative DBS-on (right) left side measurements**. f)** The same as in panel e, but for right side measurements. **g)** Stop performance as a function of UPDRS scores without and with pharmacotherapy (med-off and med-on), in preoperative (left) and postoperative DBS-on (right) left side measurements**. h)** The same as in panel g, but for right side measurements.

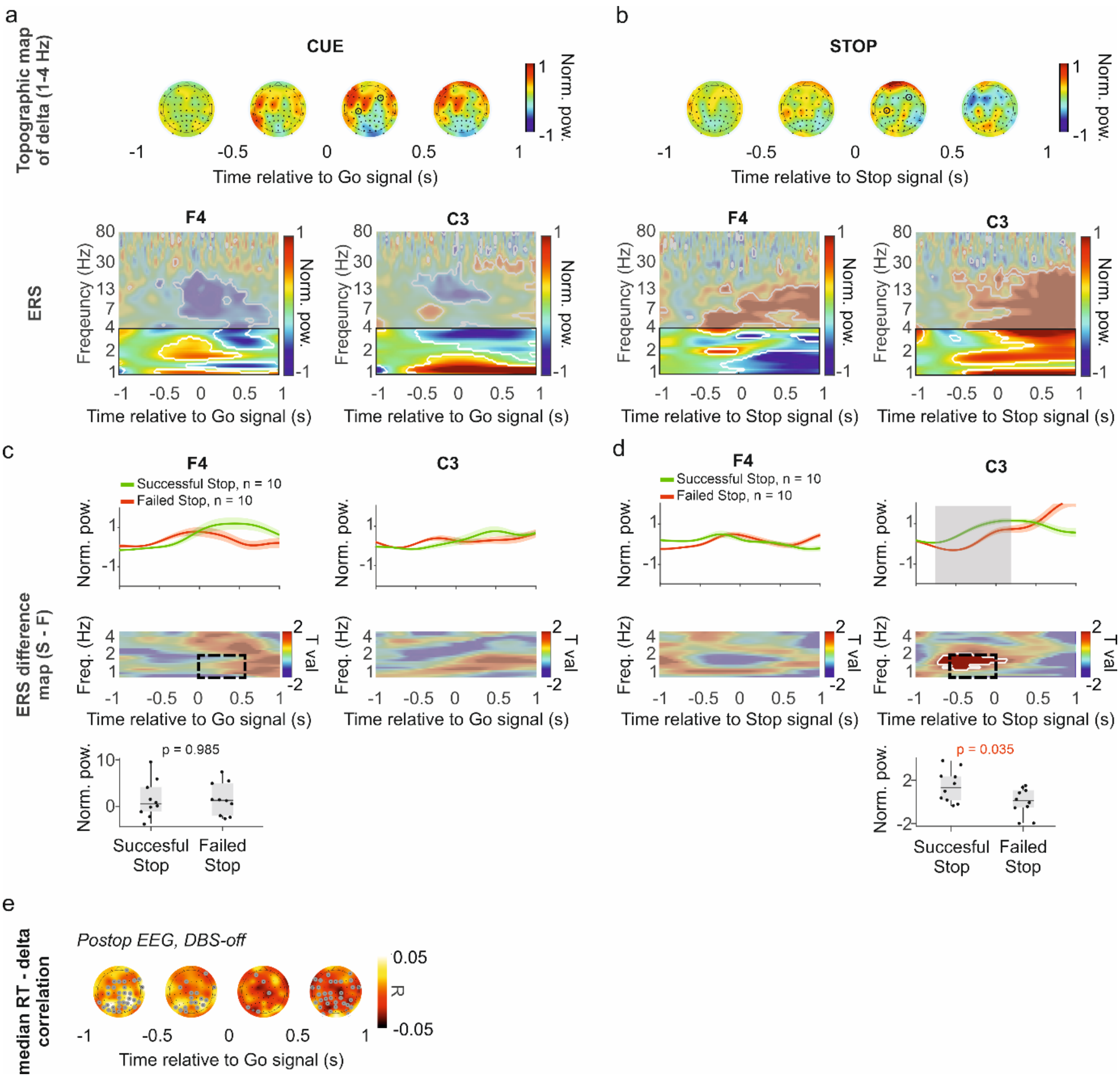


**Extended Data Figure 5. Frontal delta power changes during SSRT performance with stimulation turned on. Negative correlation of median RT and frontal delta power around go signal. a)** Top, topographical maps ('topo plots') of 60-channel EEG recordings showing delta (1-4 Hz) power derived from scalp current densities, averaged across trials and patients in 0.5 s time windows aligned to go signals. Black dots correspond to the estimated position of EEG electrodes arranged according to standard 10-20 system. Data from left side postoperative DBS-on measurements are shown. Bottom, event-related spectrograms (ERS) of selected channels (marked by black circles on the topo plots) aligned to go signals (delta frequency range highlighted). Significant changes relative to baseline are indicated by white contours (permutation test with cluster-based correction, $p < 0.05$). **b)** The same as in panel a but aligned to stop signals. **c)** Top, delta band spectral power averages aligned to go signals for successful (green) and failed (red) stop trials, averaged across trials and patients from left side postoperative DBS-on measurements, shown for the same channels as in panel a (line and shading, mean ± SE). Middle, spectrograms displaying delta power differences between successful and failed stop trials. Bottom, delta power difference between successful and failed stop trials, calculated for the time-frequency windows marked in the middle panel (black dashed box). Box-whisker plots show median, interquartile range and non-outlier range ($p = 0.985$, Mann-Whitney U-test). **d)** The same as in panel c but aligned to stop signals. Significant changes relative to baseline are indicated by white contours (permutation test with cluster-based correction, $p < 0.05$). Successful stop trials were associated with larger central delta power before the stop cue. ($p = 0.035$, Mann-

Whitney U-test). **e)** Correlation of median RT and frontal EEG delta power (postoperative recording, DBS turned off) around the go signal (Pearson's correlation coefficients). Channels with significant correlations are marked with grey (permutation statistics with false discovery rate correction, $p < 0.05$).

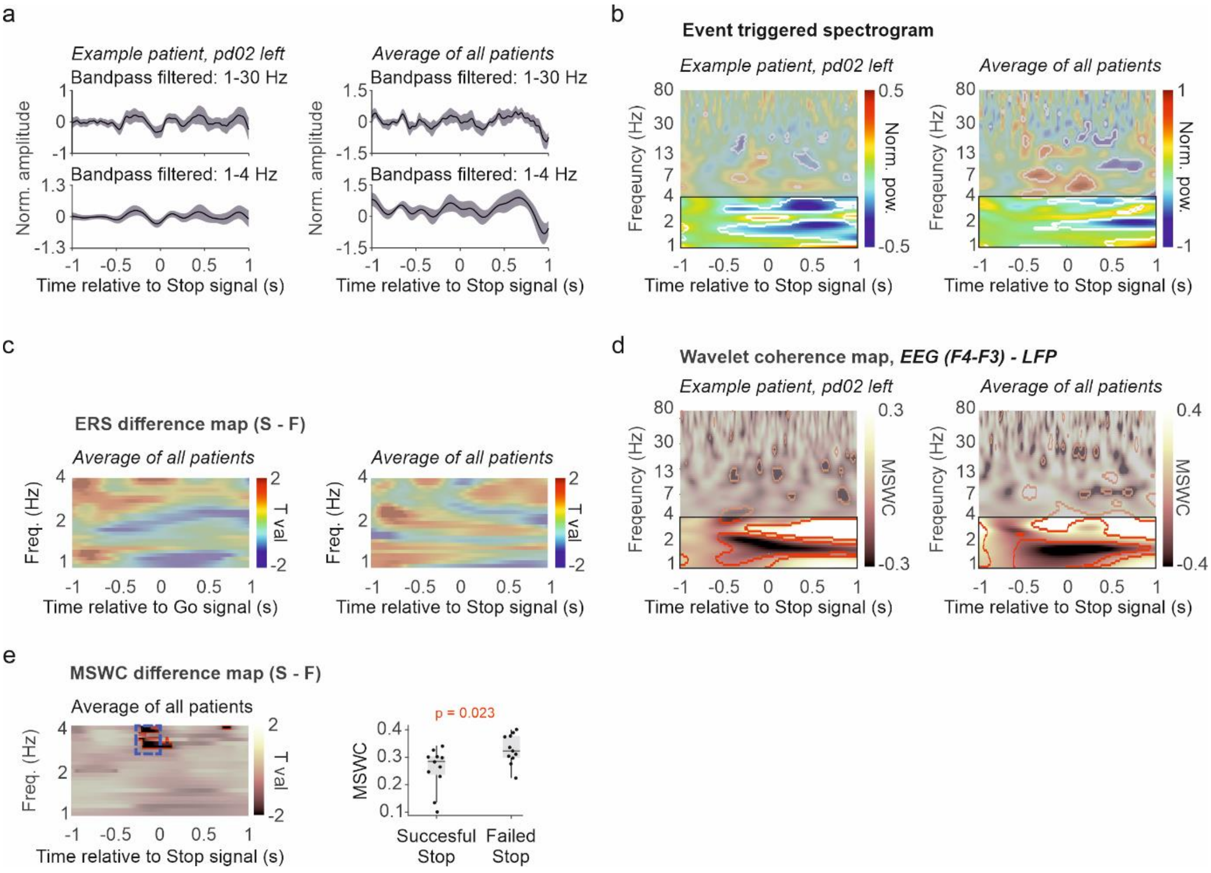


**Extended Data Figure 6. STN LFP power changes during SSRT performance. a)** Event-triggered averages (ETA) of STN local field potential (LFP) aligned to stop signals, measured during DBS implantation. Broadband (top, 1-30 Hz) and delta band (bottom, 1-4 Hz) filtered signals are shown. Shaded lines mark standard error. Left, example patient; right, average across patients. **b)** Event-related wavelet power spectrograms (ERS) relative to baseline, aligned to stop signals. Delta frequency band is highlighted; significant changes relative to baseline (-1 to -0.5 s) are marked by white contours. Left, example patient; right, average across patients. **c)** Spectrograms displaying delta power differences between successful and failed stop trials aligned to go (left) and stop signals (right), represented as paired T statistics (T val.) calculated for each time-frequency point for intraoperative STN LFP measurements. **d)** Magnitude-squared wavelet coherence (MSWC) of STN LFP and intraoperative frontal bipolar EEG aligned to stop signals. Color code indicates increase (white) and decrease (black) relative to baseline (-1 to -0.5 s). Delta frequency band is highlighted; significant changes relative to baseline are marked by white contours. Left, example patient; right, average across patients. **e)** Left, MSWC difference between successful and failed stop trials, aligned to stop signals, represented as paired T statistics (T val.) calculated for each time-frequency point, averaged across patients. White color indicates greater coherence associated with successful stop trials (permutation test with cluster-based correction, $p < 0.05$). Significant differences are marked by red contours. Right, MSWC difference between successful and failed stop trials, calculated for the time-frequency windows marked in the left panel (blue dashed box). Box-whisker plots show median, interquartile range and non-outlier range. $p = 0.023$, Mann-Whitney U-test.

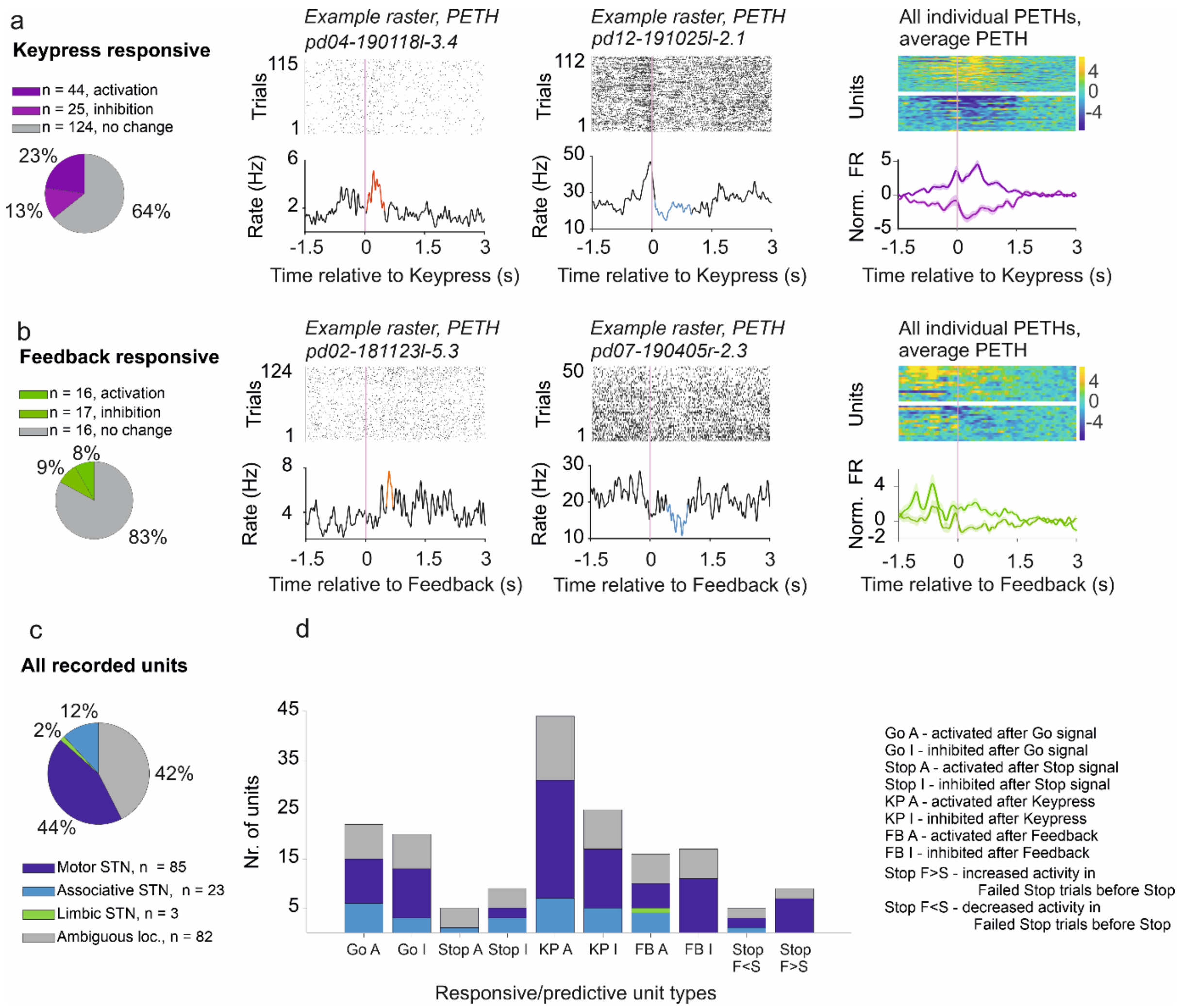


**Extended Data Figure 7. STN neurons responsive to behaviorally relevant events during SSRT. Distribution of units across STN subregions. a)** STN units responsive to Keypress. Left, proportion of Keypress-responsive units. Middle, event-aligned raster plots and peri-event time histograms (PETH) for representative activated (significantly increased activity in red) and inhibited (significantly decreased activity in blue) units. Right, heatmaps of the Z-scored PETHs of all Keypress-responsive units (yellow, firing rate increase; blue, firing rate decrease) and average PETHs of activated and inhibited units (shading, standard error). **b)** STN units responsive to Feedback. Same arrangement as in panel a. **c)** Distribution of recorded units across different STN subregions. **d)** Proportion of different types of responsive/predictive units across STN subregions.

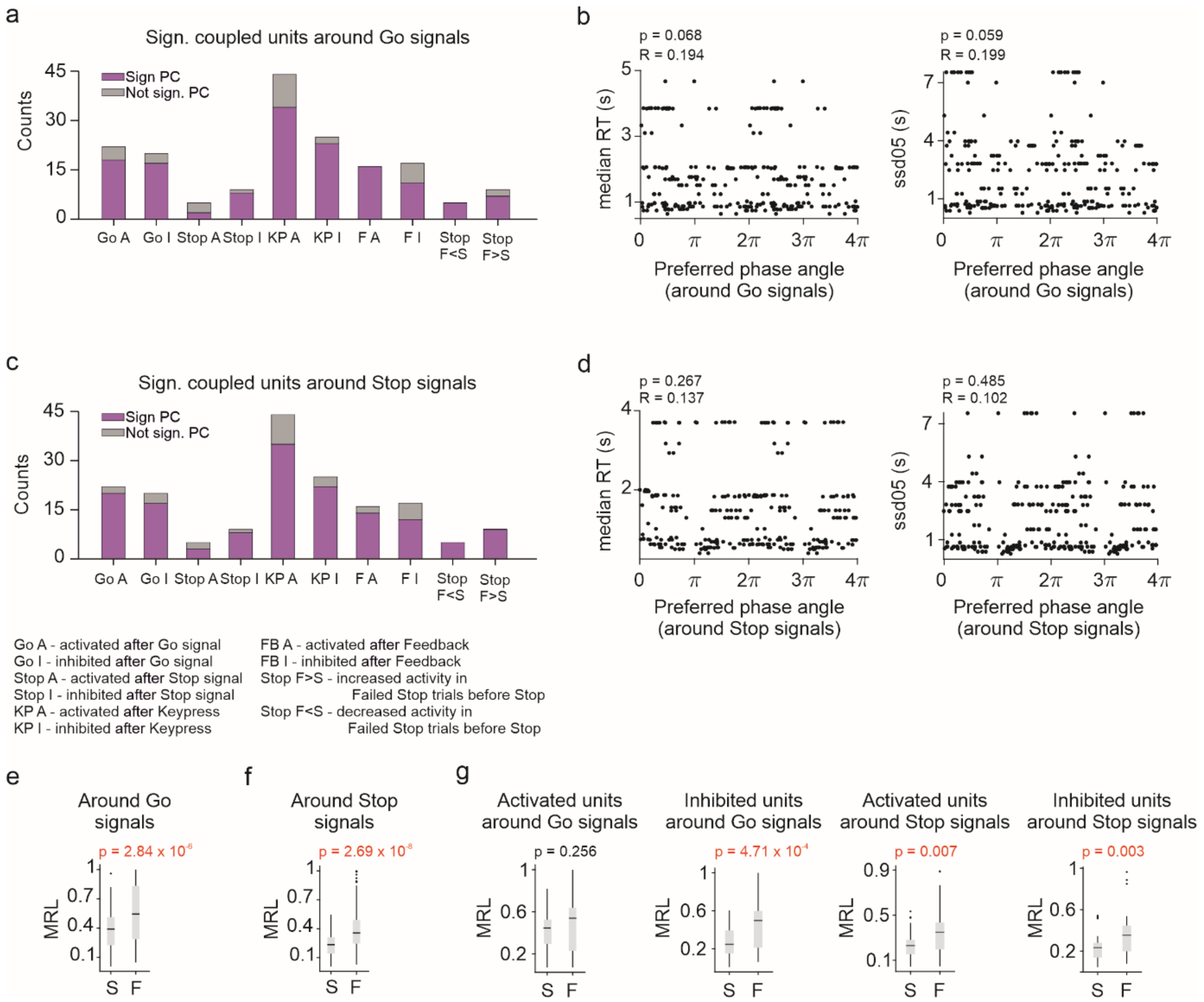


**Extended Data Figure 8. Phase coupling of STN units to local delta activity. a)** Proportion of significantly delta-coupled units among STN units responsive / predictive of SSRT task events, calculated for spikes in -1.5 s to 1.5 s time windows around go signals. **b)** Circular-linear correlation between preferred phase of delta-coupled STN units (around go signals) and SSRT task parameters: median RT (left) and SSDP0.5 (right). **c)** Same as in panel a but calculated for spikes in a -1.5 s to 1.5 s time windows around the stop signals. **d)** Same as in panel b but calculated for spikes in a -1.5 s to 1.5 s time windows around the stop signals. **e)** Comparison of coupling strength (MRL) of delta-coupled units in successful (S) and failed (F) stop trials, calculated for spikes -1.5 s to 1.5 s relative to go signals. Box-whisker plots show median, interquartile range and non-outlier range. $p = 2.84 \times 10^{-6}$, Mann-Whitney U-test. Dots mark outliers. **f)** Same as in panel e but calculated for spikes in a -1.5 s to 1.5 s time window around stop signals. **g)** Comparison of coupling strength between successful and failed stop trials in different behaviorally responsive populations (Mann-Whitney U-test).

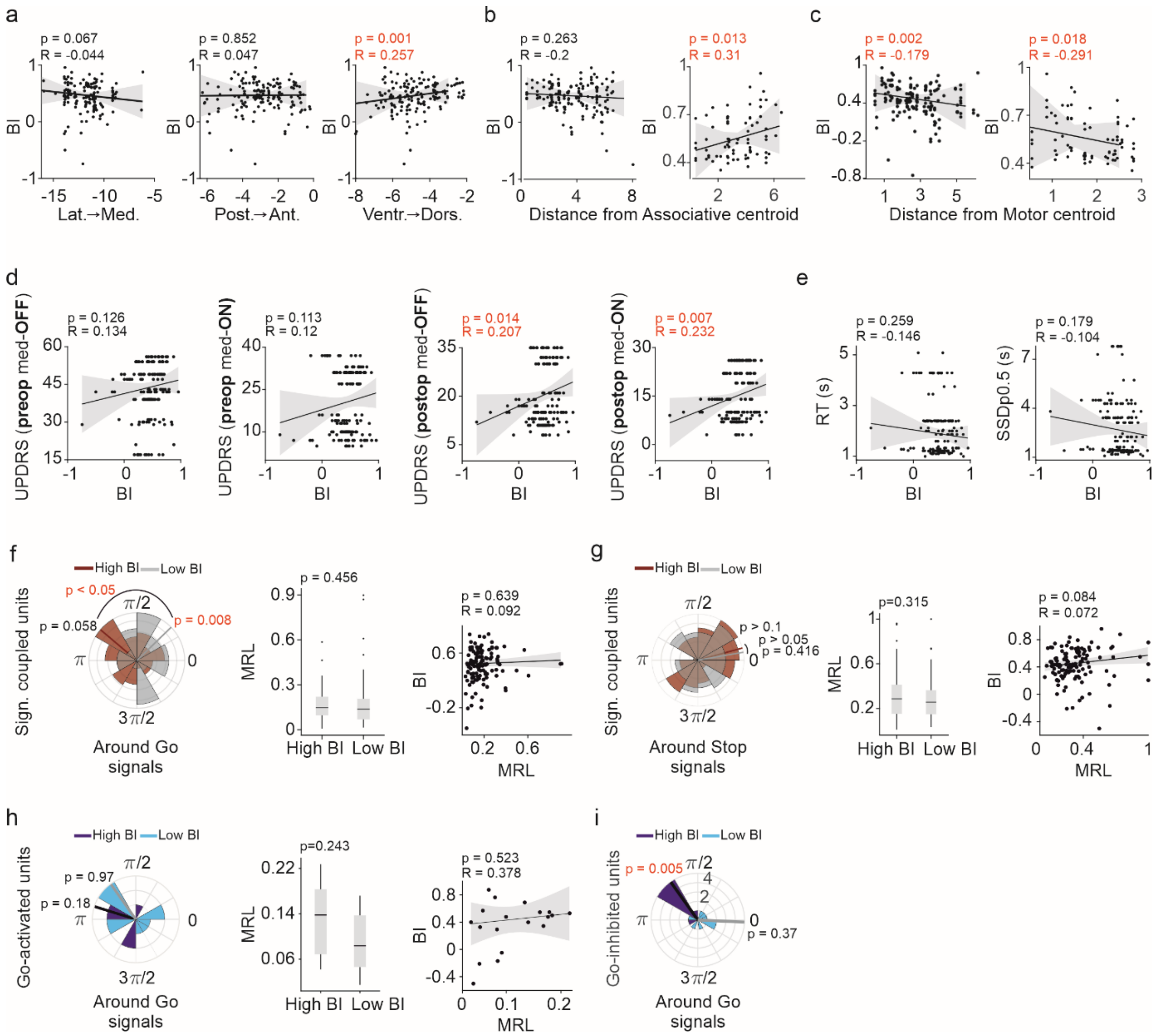


**Extended Data Figure 9. Interrelationship of SSRT task performance and STN unit bursting. a)** Correlation of bursting index (BI) with MNI coordinates of recording electrodes, considering the medio-lateral (left), postero-anterior (middle) and ventro-dorsal (right) axis. **b)** Correlation of BI with distance of electrodes from the associative centroid of the STN. Left, the correlation includes all recorded units. Right, the correlation includes only bursting units recorded on channels close to the motor centroid (closer than half of the maximal distance). **c)** Same as panel b, but distance is calculated from the motor centroid. **d)** Correlation of bursting index (BI) with UPDRS scores (pre- and postoperative, medication on and off). Lines indicate robust regression with 95% confidence intervals (grey shading). Pearson's correlation coefficients and corresponding p-values (t-test) are shown for each plot. **e)** Correlation of bursting index (BI) with SSRT task measures (RT and SSDp0.5). **f)** Left, population phase histogram of units with significant delta phase-coupling around go signals (-1.5 to 1.5 s time window), separated by median BI (grey, low BI; brown, high BI). Lines indicate mean phase. Population phase preference was tested by Rayleigh's Z-test (units with low BI, p = 0.008; units with high BI, p = 0.058). The preferred phases of low and high BI groups were significantly different (p < 0.05, two-samples Watson-test). Middle, delta phase-coupling strength (MRL) around go signals (-1.5 to 1.5 s time window) of low and high BI units were not significantly different (p = 0.456, Mann-Whitney U-test). The box-whisker plot shows median, interquartile range and non-outlier range. Dots mark outliers. Right, correlation between delta phase-coupling strength around go signals and BI of all delta-coupled units. Each dot corresponds to a unit. Line indicates robust regression with 95% confidence intervals (grey shading). Pearson's correlation coefficients and corresponding p-values (t-test) are shown. **g)** Left, population phase histogram of units with significant delta phase-coupling around stop signals (-1.5 to 1.5 s time window), separated by median BI (grey, low BI; brown, high BI). Lines indicate mean phase. Population phase preference was tested by Rayleigh's Z-test (units with low BI, $p = 0.416$; units with high BI, $p = 0.1$). The preferred phases of low and high BI groups were not significantly different ($0.05 < p < 0.1$, two-samples Watson-test). Middle, delta coupling strength of low and high BI units were not significantly different ($p = 0.315$, Mann-Whitney U-test). Right, correlation between delta coupling strength around stop signals and BI of all delta-coupled units. **h)** Same as in panel f but for units activated after the go signal, separated by median BI (light blue, low BI; dark blue, high BI). **i)** Same as left figure of panel f but for units inhibited after the go signal, separated by median BI (light blue, low BI; dark blue, high BI).

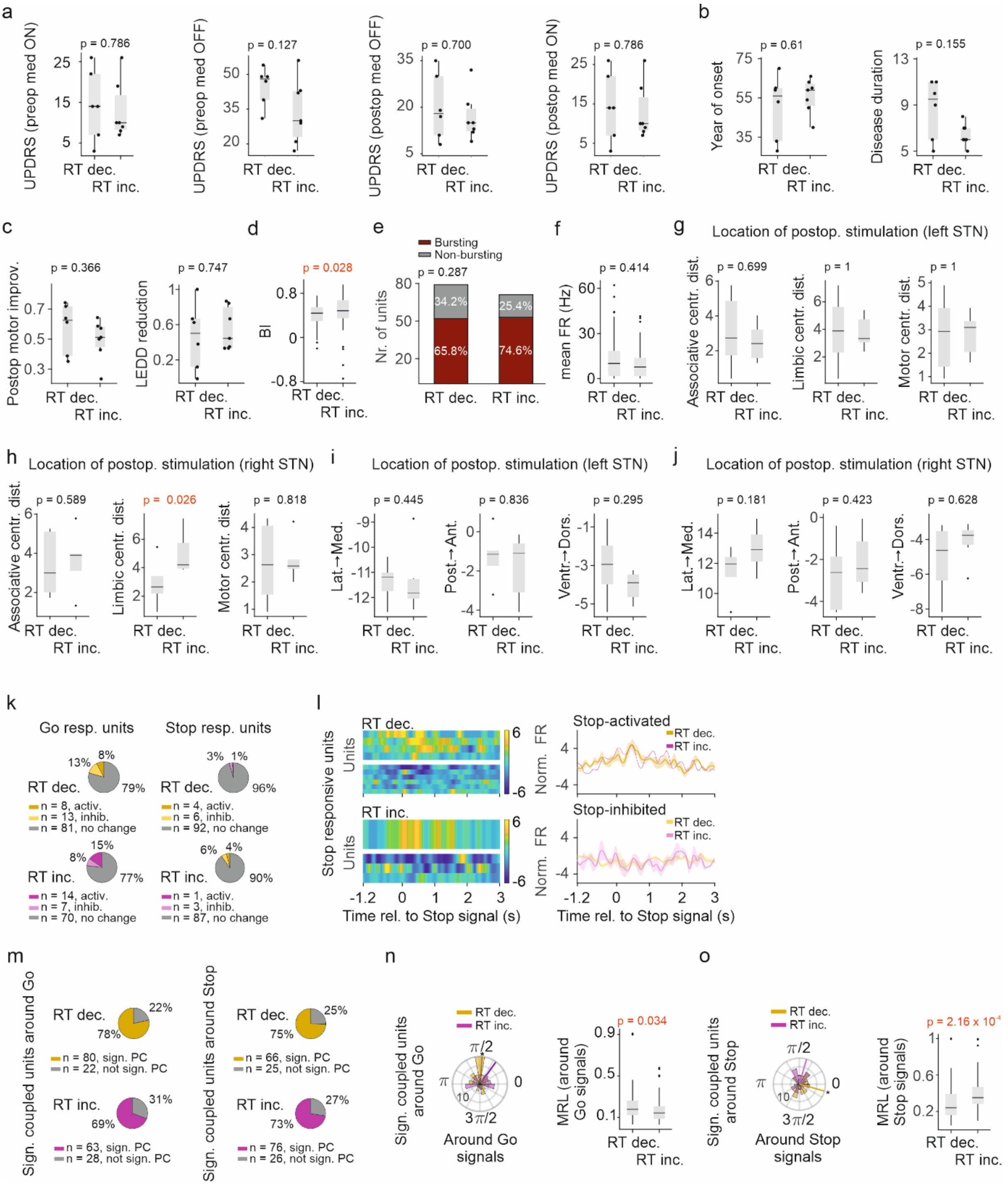


**Extended Data Figure 10. STN unit properties across patient subgroups. a)** Comparison of UPDRS scores across patients with RT decrease (RT dec.) and RT increase (RT inc.), tested with Mann-Whitney U-test. Left to right: preop UPDRS score with medication off, preop UPDRS score with medication on, postop UPDRS score with medication off and DBS turned on, postop UPDRS score with medication on and DBS turned on. Box plots represent median, interquartile range and non-outlier range (whiskers). Each dot corresponds to one patient. **b)** Comparison of year of disease onset (left) and disease duration (right) across patients with RT decrease and RT increase (tested with Mann-Whitney U-test). **c)** Comparison of postoperative motor improvement (left) and postoperative levodopa equivalent daily dose (LEDD) reduction (right) across patients with RT decrease and RT increase (tested with Mann-Whitney U-test). **d)** Comparison of BI across STN units recorded from patients with RT decrease and RT increase (tested with Mann-Whitney U-test). Box plots represent median, interquartile range, non-outlier range (whiskers) and outliers (dots). **e)** Number of bursting and non-bursting STN units recorded from patients with RT decrease and RT increase (tested with Fisher's exact test). **f)** Mean firing rate of STN units from patients with RT decrease and RT increase (compared with Mann-Whitney U-test). **g)** Distance of electrodes stimulated postoperatively in the left STN from the associative (left), limbic (middle) and motor (right) centroids from patients with RT decrease and RT increase (tested with Mann-Whitney U test). **h)** Same as in panel g but calculated for the right STN. **i)** MNI coordinates of electrodes stimulated postoperatively in the left STN compared in patients with RT decrease and RT increase, considering medio-lateral (left), antero-posterior (middle) and dorso-ventral (right) axis (tested with Mann-Whitney U test). **j)** Same as in panel i but calculated for the right STN. **k)** Left, proportion of go-responsive units recorded from patients with RT decrease vs. RT increase. Right, the same for stop-responsive units. **l)** Left, peri-event time-histogram (PETH) of stop-responsive units (color coded; yellow, activation; blue, inhibition). Right, corresponding PETH averages. No significant differences were found ($p > 0.05$, permutation test with cluster-based correction). **m)** Left, proportion of significantly delta-coupled units considering spikes in a -1.5 s to 1.5 s time window around go signals compared between patients with RT decrease vs. RT increase ($p = 0.145$, chi-square test). Right, proportion of significantly delta-coupled units considering spikes in a -1.5 s to 1.5 s time window around stop signals compared between patients with RT decrease vs. RT increase ($p = 0.755$, chi-square test). **n)** Left, population phase histogram of significantly delta-coupled units recorded from patients with RT decrease vs. RT increase (spikes in -1.5 s to 1.5 s time window around go signals were included). Lines indicate mean phase (mean angle, RT increase, 0.94 (53.9°); RT decrease, 1.5 (85.9°)). The preferred phases of STN units recorded from patients with RT decrease vs. RT increase were not significantly different (Watson's two sample test of homogeneity, $p > 0.05$). Circular uniformity was tested by Rayleigh's test (patients with RT increase, $p = 0.099$; patients with RT decrease, $p = 0.002$). Right, comparison of delta coupling strength of STN units recorded from patients with RT decrease vs. RT increase (Mann-Whitney U-test). **o)** Same as in panel n but calculated for spikes in a -1.5 s to 1.5 s time window around stop signals (mean angle, RT increase, 1.2 (68.8°), RT decrease, 6 (342.2°)). The preferred phases of STN units recorded from patients with RT decrease vs. RT increase were not significantly different (Watson's two sample test of homogeneity, $p > 0.05$). Circular uniformity was tested by Rayleigh's test (patients with RT increase, $p = 0.074$; patients with RT decrease, $p = 0.018$).

Removed to comply with medRxiv data protection policy.

**Extended Data Table 1. Summary table of study population.**
TD: tremor-dominant; AR: akinetic-rigid; MX: mixed.

Removed to comply with medRxiv data protection policy.

**Extended Data Table 2. Summary table of STN-DBS stimulation parameters.**

| Type of units compared with other behaviorally relevant units | P value |
|---|---|
| Go activated | 0.0691 |
| Go inhibited | 1.0000 |
| Go activated + Go inhibited | 0.0752 |
| Stop activated | 0.2121 |
| Stop inhibited | 0.0601 |
| Stop activated + Stop inhibited | 0.0158 |
| Stop predictive type 1 | 0.3303 |
| Stop predictive type 2 | 0.5170 |
| Stop predictive type 1+2 | 0.4914 |
| Key press activated | 1.0000 |
| Key press inhibited | 0.4914 |
| Key press activated + Key press inhibited | 0.3177 |
| Feedback activated | 0.0868 |
| Feedback inhibited | 0.1024 |
| Feedback activated + Feedback inhibited | 1.0000 |

**Extended Data Table 3. Testing localization bias of behaviorally responsive / predictive unis in STN subregions.** The proportion of units localized in the associative and motor STN subregions was compared with Fisher's exact test.
